# Multilevel and Quasi Monte Carlo methods for the calculation of the Expected Value of Partial Perfect Information

**DOI:** 10.1101/2021.03.30.21254626

**Authors:** Wei Fang, Zhenru Wang, Michael B. Giles, Chris H. Jackson, Nicky J. Welton, Christophe Andrieu, Howard Thom

## Abstract

The expected value of partial perfect information (EVPPI) provides an upper bound on the value of collecting further evidence on a set of inputs to a cost-effectiveness decision model. Standard Monte Carlo (MC) estimation of EVPPI is computationally expensive as it requires nested simulation. Alternatives based on regression approximations to the model have been developed, but are not practicable when the number of uncertain parameters of interest is large and when parameter estimates are highly correlated. The error associated with the regression approximation is difficult to determine, while MC allows the bias and precision to be controlled. In this paper, we explore the potential of Quasi Monte-Carlo (QMC) and Multilevel Monte-Carlo (MLMC) estimation to reduce computational cost of estimating EVPPI by reducing the variance compared with MC, while preserving accuracy. In this paper, we develop methods to apply QMC and MLMC to EVPPI, addressing particular challenges that arise where Markov Chain Monte Carlo (MCMC) has been used to estimate input parameter distributions. We illustrate the methods using a two examples: a simplified decision tree model for treatments for depression, and a complex Markov model for treatments to prevent stroke in atrial fibrillation, both of which use MCMC inputs. We compare the performance of QMC and MLMC with MC and the approximation techniques of Generalised Additive Model regression (GAM), Gaussian process regression (GP), and Integrated Nested Laplace Approximations (INLA-GP). We found QMC and MLMC to offer substantial computational savings when parameter sets are large and correlated, and when the EVPPI is large. We also find GP and INLA-GP to be biased in those situations, while GAM cannot estimate EVPPI for large parameter sets.

## 1 Introduction

Cost-effectiveness analysis is used to compare the costs and benefits of medical interventions, which are often combined as a net monetary benefit (NB)[1]. Such analyses are internationally adopted by decision makers and Health Technology Assessment (HTA) agencies, including the National Institute for Health and Care Excellence (NICE) in the UK, the Institute for Clinical Effectiveness Review in the US, the Canadian Agency for Drugs and Technology in Health (CADTH) in Canada, and the Pharmaceutical Benefits Advisory Committee (PBAC) in Australia. Costs and effects of interventions can be estimated using trial-based analysis or extrapolated over patient lifetimes using model-based decision analysis [2]. Examples of decision models include decision trees, cohort Markov models, and individual patient microsimulations [3]. These models estimate the net benefit of interventions as a function of input parameters such as treatment effectiveness, drug costs or quality of life following an intervention. This function will be referred to as the net benefit (NB) function. Because the model parameters are estimated from data, we are uncertain about the true parameter values (due to imperfect information on them). This uncertainty in model parameters is propagated through the model to give our uncertainty in the true costs and effects. This in turn leads to a quantification of our uncertainty around the optimal treatment recommendation, which is known as decision uncertainty arising from imperfect information on input parameters. Expected value of perfect information (EVPI) is the expected improvement in decision making, often valued on the monetary scale, from gaining perfect information on all parameters [4, 5]. Expected value of partial perfect information (EVPPI) is the value of gaining perfect information on a subset of the parameters [6]. These quantities have the potential to guide research funding, as studies costing more than the EVPI and EVPPI will not be cost-effective but those costing less may be cost-effective and this can be explored using the Expected Value of Sample Information (EVSI). EVPPI is also a useful sensitivity analysis as it can highlight parameters to which the decision is most sensitive.

Estimating EVPPI requires nesting one Monte Carlo simulation over the subset of parameters on which further research is being considered within a second Monte Carlo simulation of the remaining parameters. This procedure is termed nested Monte Carlo and requires evaluation of the NB function for all samples. EVPPI calculations can be computationally intensive, especially for economic models which involve individual level simulation or Markov models with large numbers of states, and incur significant computational cost to evaluate the NB functions. As a result, standard nested Monte Carlo simulation often fails since it requires an impractical number of samples to get a reasonably precise estimate. In addition, estimation based on the standard nested Monte Carlo method is biased [6, 7]. Estimation of this bias is computationally expensive as it requires comparison of EVPPI estimates based on different numbers of samples [6, 8]. Much recent effort has aimed to reduce the computational burden of EVPPI [7] through approximating the conditional expected NB functions, and thus replacing one of the Monte Carlo samples from nested Monte Carlo, by some functions which are less computationally intensive to evaluate. Success has been found through linear approximations to the conditional expected NB functions or exact algebraic solutions of the EVPPI, but these methods are specific to each model design and are not always appropriate, particularly for highly complex and nonlinear model structures [9]. Meta-modelling through generalised additive models, Gaussian processes and Integrated Nested Laplace Approximations (INLA-GP), is an elegant and general approach to reducing the computational burden of EVPPI [10, 11, 12]. GP and GAM methods fit a regression model of net benefit on the input parameters to then estimate the conditional expected NB, thus removing the need for nested simulation. The INLA-GP method fits a 2D Gaussian process to a dimension-reduced sample of the model parameters and model outputs. These have been implemented in user friendly online tools and software packages [13, 14]. However, all these methods are based on approximating the conditional expected NB function, incurring a bias which is difficult to quantify. A prohibitively large number of samples may also be required to determine a sufficiently well-fitting regression function.

Cost-effectiveness decision models should reflect all the available relevant evidence, to fully capture decision uncertainty. For model parameters where there are multiple evidence sources available, evidence synthesis methods are used to pool the results, often using Bayesian inference evaluated using Monte Carlo Markov chain simulation [1]. Relative treatment effects, for example, are commonly estimated using Bayesian network meta-analysis (NMA), which delivers a joint distribution for multiple treatment effects that are not available in closed form but instead represented by samples from an MCMC simulation. This poses two challenges for EVPPI calculation. Firstly it may require a very large number of samples to characterise the posterior distribution, for example if correlations between parameters impede mixing of the sampler. In addition to needing a large number of samples, it can also be computationally expensive to generate each sample from the posterior, as for the NMA used in the directly acting oral anticoagulants (DOACs) for prevention of stroke in atrial fibrillation Markov model [15, 16, 17, 18]. This can make nested Monte Carlo, or the generation of a sufficient number of samples to determine the regression function for GAM or GP methods, impractical. A further challenge with MCMC is that it is difficult, particularly using off-the-shelf general purpose Gibbs samplers, such as OpenBUGS [19], to generate the conditional distributions needed for EVPPI estimation by standard nested Monte Carlo. This motivates us to explore new computational methods for EVPPI.

In this paper, instead of approximating the conditional expected NB function, we introduce two different Monte Carlo methods to reduce the number of samples and NB function evaluations needed for the same accuracy as the standard nested Monte Carlo method: Multilevel Monte Carlo (MLMC) and Quasi Monte Carlo (QMC). These Monte Carlo methods are unrelated to each other but we explore them in parallel.

The multilevel Monte Carlo (MLMC) method, introduced by Giles [20, 21], has been successfully applied to many research fields, such as financial mathematics, mathematical biology and uncertainty quantification. The first key idea of MLMC is to create a series of estimators for the quantity of interest, such as EVPPI, which are increasing in accuracy and increasing in computational cost. The first term, which is the lowest ‘level’, is the least accurate and computationally least intensive, while the last term, or highest level, is the most accurate and most expensive. All these terms have similar variance. By careful construction, consecutive estimators in the sequence are designed to be highly correlated. This gives the second key idea, which is that the differences between consecutive terms have much lower variance than the individual terms and thus require fewer total samples to estimate. A combined estimator for the quantity of interest can then be formed by adding the lowest level estimator to a sum of the differences of consecutive terms, which is a sum over the levels of MLMC and has the highest accuracy. As this is formed of terms that have much lower variance, the total number of samples needed for estimation can be much reduced from that needed for standard (i.e. non-multilevel) nested Monte Carlo [21]. Goda and others have used MLMC to construct an estimator of the EVPPI that has lower cost than standard nested Monte Carlo (MC) [22, 23]. Technical details of MLMC for EVPPI are provided in Section 2.2 of this paper. MLMC for EVPPI offers greatest computational savings over MC when there are correlations between model parameters. This is because greater correlation requires more inner samples, and thus deeper levels of MLMC, which leads to proportionately greater computational savings over MC. An additional benefit of MLMC for EVPPI is that it can be used to estimate the bias of EVPPI, and indeed EVPI, and this estimate can be used to form an unbiased estimator of EVPPI [22, 24]. Inference on EVPPI often focusses on the point estimate and disregards uncertainty in the estimate, which could have an impact on trial funding decisions. An estimate of both the accuracy and precision of the estimator would therefore be useful, beyond the need to form an unbiased estimator. Previous work has applied MLMC for EVPPI to only simple models and has not considered the case where the model input parameters have been estimated using Bayesian methods, and uncertainty in the parameters is represented by MCMC simulations rather than a closed form distribution.

Standard Monte Carlo simulation uses pseudo-random number generators that attempt to mimic truly random sequences [25]. However, with truly random sequences, the generated sequences tend to include clusters of points separated by large gaps. An example is shown in Figure 2(c), which is a standard Monte Carlo sample from a bivariate uniform distribution. These clusters and gaps tend to reduce the efficiency of estimators based on the sample. Quasi Monte Carlo (QMC) [26], is an alternative to standard Monte Carlo simulation that instead of using pseudo-random number generators, uses quasi-random sequences which have been specially designed to avoid bunching and gaps in the sampling space (known as “low discrepancy” sequences). This results in an estimator with a lower variance, which in turn reduces the number of necessary samples and computational cost. However, QMC has not yet been applied to EVPPI.

**Figure 1:**
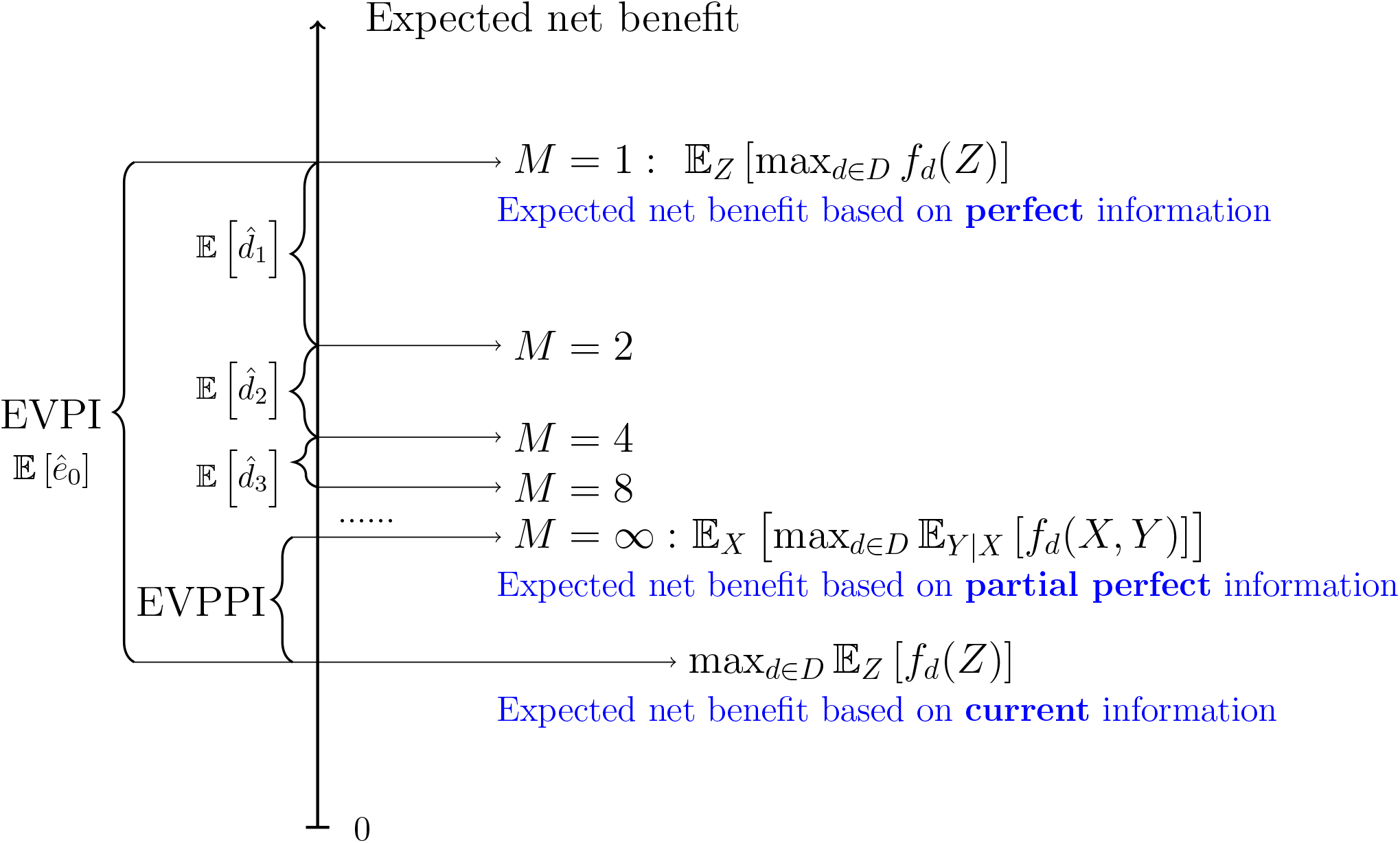
Illustration of multilevel Monte Carlo (MLMC) estimation of EVPPI. The horizontal lines represent estimates of the expected net benefit under partial perfect information, using *𝓁* levels and *M* = 2^*𝓁*^ inner samples. The MLMC estimate of EVPPI is the difference between the level *𝓁* estimate and the expected net benefit under current information. For *𝓁* = 0 this is the EVPI, while it converges to the true EVPPI as *𝓁* → ∞, or equivalently, as an increasing number of bias reduction terms 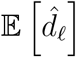 are added to the *𝓁* = 0 estimator.

**Figure 2:**
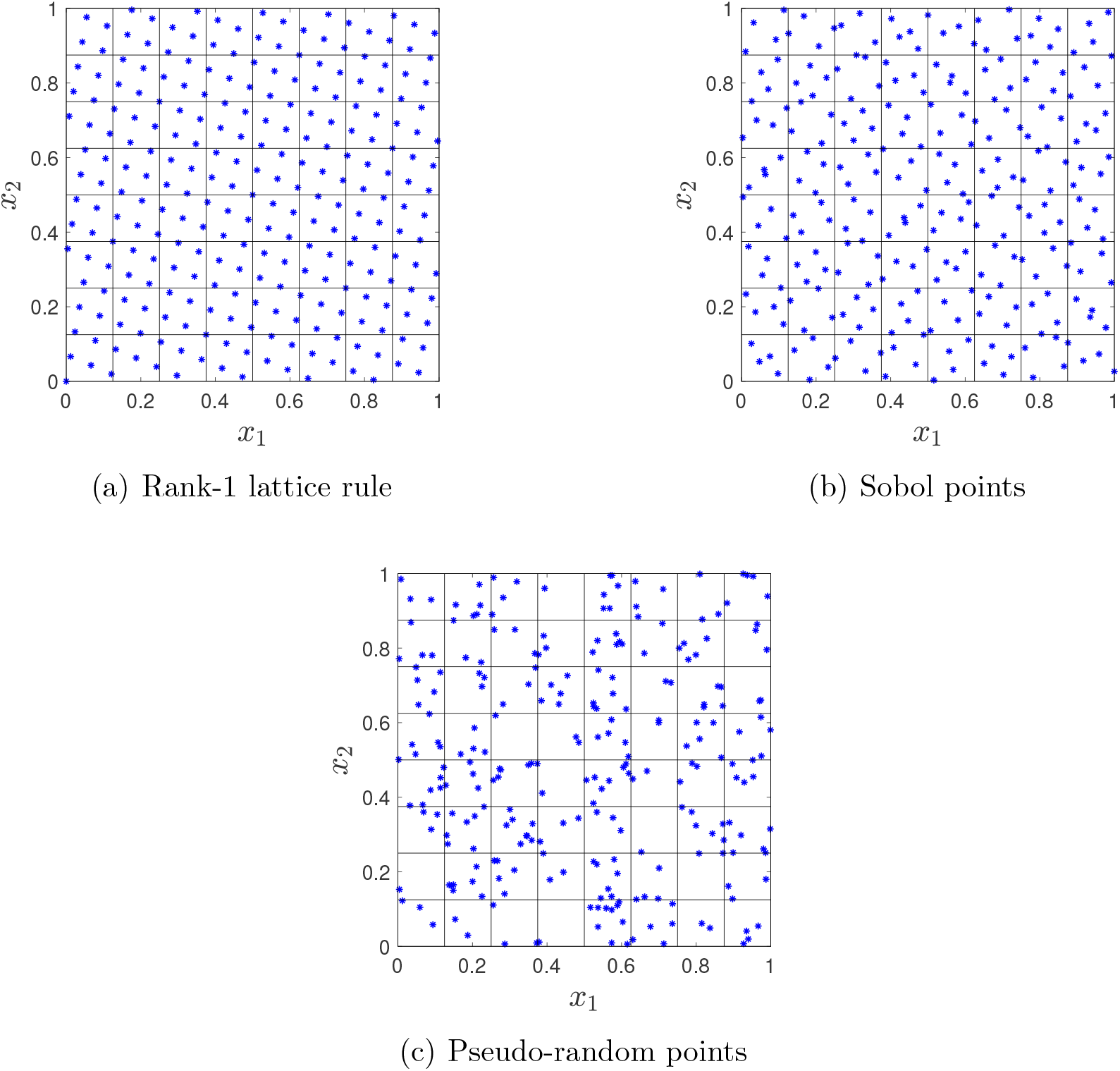
Generating points: QMC vs. MC.

Both MLMC and QMC are variance reduction methods that allow accurate estimation of the expectations with fewer samples than nested MC. Many other variance reduction methods are available but out of the scope of this paper, e.g. control variables, importance sampling, stratified sampling and Latin hypercube, see [27] for more comprehensive description of these methods.

This paper develops a QMC approach, and extends an existing MLMC approach, to efficient EVPPI estimation. In Section 2 we begin with a discussion of the properties of Monte Carlo nested simulation for EVPI and EVPPI. How MLMC and QMC can reduce the number of samples needed and the extension of MLMC and QMC to MCMC samples are provided in Section 3. Section 4 presents an application to an artificial, but realistic, decision tree depression model with MCMC input parameters. Section 5 presents an application to the published and highly complex DOACs for prevention of stroke in atrial fibrillation Markov model [16, 17, 18]. Further discussions and conclusions are provided in Section 6. In the Appendix, in addition to detailed experiment results, we also show how MLMC and QMC can be applied to value of information analysis, through a step-by-step explanation of the method and the provision of code examples.

## 2 Methods

### 2.1 Standard nested Monte Carlo estimation of EVPI & EVPPI

Suppose our model is a function of input parameters *Z*, and the net benefit in monetary units for each decision option d is given by *f*_*d*_. Then assuming the decision maker is risk neutral and rationale, the optimal decision option is that which the expected net monetary benefit:

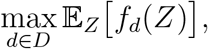

where 𝔼_*Z*_ is the expectation with respect to *Z*. If all uncertainty is eliminated in the parameter inputs, then we can choose the best decision and get monetary benefit max_*d*∈*D*_ *f*_*d*_(*Z*). Taking an expectation over the possible realizations of *Z* gives the expected monetary value based on perfect information

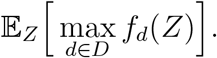

Then, we define EVPI as the extra monetary value expected from learning the true value of *Z*:

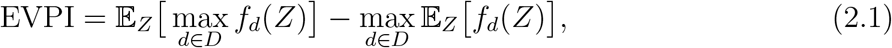

which indicates to the decision maker whether the optimal decision option is sensitive to the uncertainty in the model inputs, and whether it is potentially cost-effective to fund new research on *Z*.

It may be that we are only interested in eliminating uncertainty on a subset *X* of model input parameters, and the remaining subset *Y* of parameters are still uncertain, where *Z* = (*X, Y*). Then given perfect information on *X*, the optimal decision option is that which maximises the conditional expected net monetary benefit where the expectation is over *Y* conditional on *X*

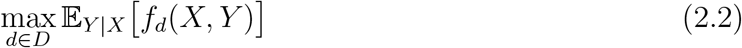

Taking an expectation over the possible realizations of *X* gives the expected monetary value

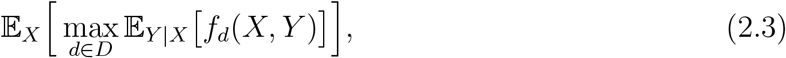

Similarly, we define the EVPPI as the increased value expected from gaining perfect information about *X*:

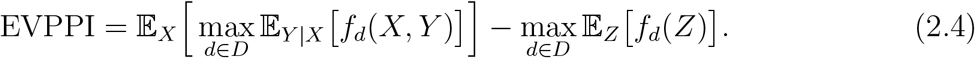

In most practical cases, there is no closed form solution to the EVPI and EVPPI and we must turn to numerical methods. One straightforward way is the nested Monte Carlo (MC) method which approximates the expectation as an average of samples. Assuming we can sample from the distribution of *Z*, we can generate *N* independent samples *Z*^(1)^, *Z*^(2)^, …, *Z*^(*N*)^ and approximate the EVPI by

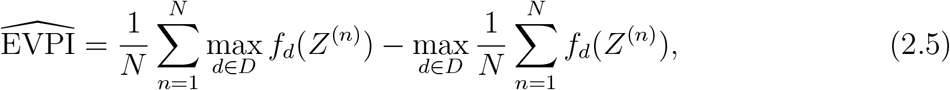

which is biased downward due to the positive bias of the second term [8].

We can form a similar approximation for EVPPI if we can sample from *X*, and from *Y* given *X*. We first generate *N* samples *X*^(1)^, *X*^(2)^, …, *X*^(*N*)^, and for each *X*^(*n*)^, we then generate *M* samples of *Y* ^(*n,m*)^ according to the conditional distribution based on *X*^(*n*)^. Finally we approximate the EVPPI by

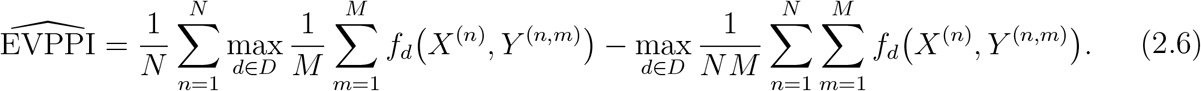

The *N* samples *X*^(*n*)^ are referred to as the ‘outer’ samples while the *M* samples of *Y* ^(*n,m*)^, the summation over which is nested inside the summation over the outer samples, are referred to as the ‘inner’ samples. Due to Jensen’s inequality, both terms in estimator (2.6) have positive bias. The estimator is therefore biased, but it is difficult to conclude whether the estimator is biased upward or downward [8].

Both the bias and variance of the estimator are important. Bias relates to accuracy of the estimate, whereas the variance relates to how precise the estimate is. A precise biased estimate can be very misleading, because it gives confidence but in the wrong thing. It’s therefore good practice to report both, and as noted the bias estimate can help obtain an unbiased estimate. In general we want to find minimum variance unbiased estimators if possible. Therefore, we consider the mean square error (MSE) of the EVPPI which is defined as

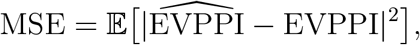

and can be decomposed into two parts,

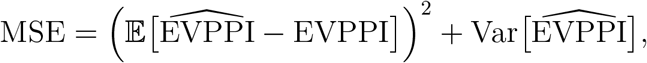

i.e., this is the square of the bias plus the variance of the estimator. For all the numerical methods, a prescribed MSE *ε*^2^ can be achieved by bounding the bias by *ε/*2 and the variance by 3*ε*^2^*/*4, since *MSE* = *Bias*^2^ +*V ar* = (*ε/*2)^2^ +3*ε*^2^*/*4 = *ε*^2^; this choice is somewhat arbitrary but we found in numerical experiments that the split has little impact on results. For a square root mean squared error (RMSE) *ε*, the total number of samples required by standard nested Monte Carlo method is of the order of *ε*^−3^ [21]. This is because the number of outer samples to bound the variance is the inverse of the desired variance (i.e. the order of *ε*^−2^) and the number of inner samples, per outer sample, to bound the bias is the inverse of the desired bias (i.e. the order of *ε*^−1^). As both the variance and bias must be bounded this gives the total order of inverse cube of the desired RMSE (i.e. *ε*^−3^).

However, in practice, nested MC is computationally expensive. In the following two sections, we introduce two advanced Monte Carlo methods to improve the computational efficiency and determine an appropriate number of samples to achieve a prescribed bias, and obtain corresponding credible intervals systematically.

### 2.2 Multilevel Monte Carlo estimation of EVPPI

We follow the recently published approach of Giles & Goda 2018 [23] to apply MLMC to the estimation of EVPPI. As explained in the Introduction, the first key idea of MLMC is to create a series of estimators of the quantity of interest, in our case EVPPI, which are increasing in accuracy, and increasing in computational cost. These estimators are carefully constructed to ensure that consecutive terms are correlated; this gives the second key idea that differences between consecutive terms have low variance, and therefore require fewer total samples to estimate compared to standard Monte Carlo. To illustrate how the MLMC method works, we first consider a simple example; full details on how to construct the necessary estimators for EVPPI will follow below.

Firstly consider two crude estimators for EVPPI, labelled *ê*_0_(*N*) and *ê*_1_(*N*), each defined as the estimator (2.6) based on *N* outer samples of *X* but with *M* = 1 and *M* = 2 inner samples of *Y* respectively, and each rewritten in the form of an average,

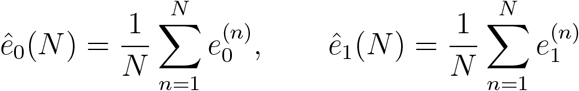

Where

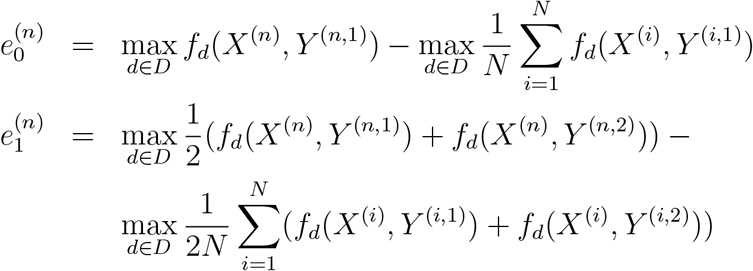

*ê*_0_(*N*) is more biased (essentially it is the estimator (2.5) for EVPI), but requires half as many samples of *Y* to achieve the same precision as the less biased estimator *ê*_1_(*N*). We can then construct a new estimator

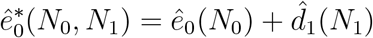

with the same degree of bias as *ê*_1_(*N*), by adding an estimator of the *bias reduction*, 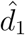, to the original biased estimator *ê*_0_. The bias reduction estimator is defined as the difference

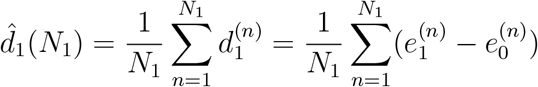

Then if each term 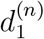 is calculated from a single sample *Y* ^(*n*,1)^, rather than using a pair of different samples of *Y* ^(*n*,1)^ for 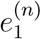 and 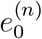, and using a single sample of *X*^(*n*)^, the *variance* of 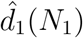 is lower. Consequently, half the number of samples are required for the new “bias-reduced” EVPPI estimator 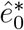 to achieve a variance similar to the original “less-biased” estimator *ê*_1_. The sizes *N*_0_, *N*_1_ of the samples used to obtain each term in the new estimator can be tuned to achieve the desired balance of variance and computational cost.

MLMC is a natural extension of this principle. A sequence of estimators *ê*_0_(*N*_0_), *ê*_1_(*N*_1_), *ê*_2_(*N*_2_), … is constructed, with decreasing bias, but also with increasing computational cost for the same precision. The *𝓁*th term in the sequence *ê*_*𝓁*_(*N*_*l*_) is again defined by the standard Monte Carlo estimator (2.6), with *N*_*𝓁*_ outer samples and *M* = 2^*𝓁*^ inner samples. Each level can have a different number of outer samples *N*_*𝓁*_ but in practice we use the same number *N* for each. The MLMC estimator of EVPPI is then constructed by starting with the most-biased estimator, and adding a sequence of bias-reduction terms,

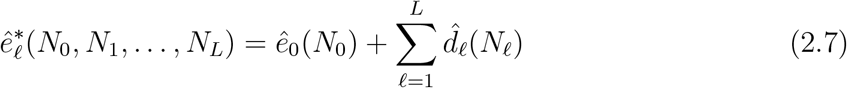

where the *𝓁*th bias reduction term is 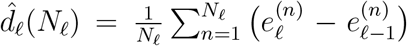, and 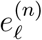 is defined as before so that the standard (non-MLMC) Monte Carlo estimator can be expressed as an average 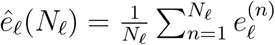 (as illustrated in Figure 1). This MLMC EVPPI estimator has the same expectation (or degree of bias) as the most expensive of the Monte Carlo estimators in this sequence, *ê*_*L*_(*N*_*L*_). And again, if the pair of components 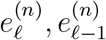 in each term of the bias reduction estimator are evaluated using the same sample *Y*, then we will require half the number of samples to achieve the same precision as an estimator built from two independent samples of *Y*. The computational savings, compared to full Monte Carlo, accumulate as the bias reduction terms are added. The consequence is that the MLMC EVPPI estimator requires fewer samples to achieve the same degree of precision as the Monte Carlo estimator *ê*_*L*_(*N*_*L*_), without affecting the bias.

As shown in [23], given a required RMSE of *ε*, the total number of samples required is of an order of magnitude of *ε*^−3^ for the standard Monte Carlo method, but *ε*^−2^ for the MLMC method at best, as long as the number of inner samples for *ê*_*𝓁*_(*N*_*𝓁*_) is set to 2^*𝓁*^. As *L* is increased, the bias of the resulting EVPPI estimator reduces, so that it converges to the true EVPPI with diminishing returns (as illustrated in Figure 1).

The number of levels *L*, and the number of outer samples *N*_*𝓁*_ in each level, can be tuned so that the MLMC estimator achieves a specific balance of bias, precision and computational speed, as outlined in Appendix 7.1.1 along with two other refinements to improve performance of the algorithm.

### 2.3 Quasi Monte Carlo estimation of EVPPI

As explained in the Introduction, standard Monte Carlo samples are “pseudo-random”, i.e. random samples from a distribution of interest, that are generated deterministically by a computer, but statistically indistinguishable from truly random samples. Quasi-random samples, on the other hand, while not statistically random, can be used to approximate the distribution of interest with fewer samples required for the same level of precision. Some examples of quasi-random samples are in Figure 2(a) and 2(b), for a bivariate uniform distribution. These can be seen to cover the sampling space more evenly than a standard Monte Carlo sample (Figure 2(c)).

For example, given a scalar random variable *X* whose cumulative distribution function Φ is known, we can generate random samples *X*^(*n*)^ of *X* by sampling *U* ^(*n*)^ from a standard uniform distribution and setting *X*^(*n*)^ = Φ^−1^(*U* ^(*n*)^). Then we can estimate, e.g. 𝔼[*X*], from a sample of size *n* by 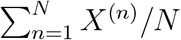. In standard Monte Carlo, *U* ^(*n*)^ are chosen randomly. In QMC, we generate *low-discrepancy* sequences, which are points distributed over [0, 1] more evenly. This approach generalises to *s*-dimensional random variables *X*, by generating *U* ^(*n*)^ distributed uniformly over the [0, 1]^*s*^ hypercube (after applying a Cholesky decomposition if the *X* are correlated, see e.g. [3]).

Two common approaches for generating low-discrepancy sequences are *rank-1 lattice* rules (illustrated in Figure 2(a)) and *Sobol* sequences (Figure 2(b)).

- Rank-1 lattice rules generate samples as 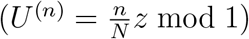, where *z* is an *s*-dimensional vector whose components are integers which share no positive integer divisors, other than 1, with the sample size *N*. The fraction 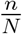 is multiplied by each element of *z* and the mod1 operation keeps only the fractional part of 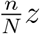. For example, if *X* consists of one scalar parameter, this would produce an evenly spaced sequence on [0,1], equivalent to equally-spaced quantiles of the distribution of *X*. For two (or more) random variables, this produces an evenly spaced “rotated” grid of points which will capture their correlation better than a simple grid (Figure 2(a)). Methods for constructing such a vector are explained in [28].
- Sobol sequences *U* ^(*n*)^ have the property that for small dimensions *s <* 40, the subsequence 2^*m*^ ≤ *n* ≤ 2^*m*+1^ of length 2^*m*^ +1, for any *m* ≥ *s*, has precisely 2^*m*−*s*^ points in each of the cubes of volume 2^−*s*^ formed by bisecting the unit hypercube in each dimension. These can be constructed as explained in [29]. For example, cutting it into halves in any dimension, each has 2^*m*−1^ points; cutting it into quarters in any dimension, each has 2^*m*−2^ points. We chose to use Sobol sequences as they are generated easily using the R package ‘randtoolbox’, illsutrated in our supplementary code [30].

In the bivariate uniform example, using a rank-1 lattice method (Figure 2(a)) there are exactly 4 points in each small square, and using a Sobol sequence (Figure 2(b)), there are roughly 3-5 points in each small square. Thus the sample space is covered more evenly than in the Monte Carlo method (Figure 2(c)).

A QMC estimator of EVPPI is defined by substituting the QMC sample Φ^−1^(*U* ^(*n*)^) for the standard Monte Carlo sample *X*^(*n*)^ in Equation (2.6). However, as the QMC sample is deterministic, it is difficult to quantify uncertainty in the estimate arising from the limited sample size. To obtain a credible interval, we instead use *randomised* QMC. This procedure generates *K* sets of QMC points {*U* ^(*k,n*)^}_1≤*n*≤*N*_ for *k* = 1, 2, …, *K* as follows, where a choice of 8 to 32 for *K* has been empirically demonstrated as sufficient [31]. We do not want to use a too large *K* as the larger *K* is, the more precision we have to sacrifice in order to obtain the credible interval.

To do this for Sobol sequences, we perform digital scrambling using a bitwise exclusive-or operation. It maintains the low discrepancy by implementing a uniform random permutation of 0 and 1 for the binary expression of each sample *U* ^(*k,n*)^ [29]. This is also implemented in the ‘randtoolbox’ R package and illsutrated in our supplementary code [30].

Then the randomised QMC estimator of EVPPI is defined by substituting Φ^−1^(*U* ^(*k,n*)^) for the standard Monte Carlo sample *X*^(*n*)^ and then averaging over random samples *k*

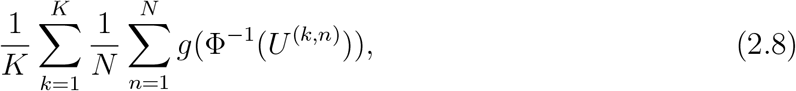

For the estimation of EVPPI, in most practical cases, the random variables *X* have standard distributions whose inverse cumulative distribution functions Φ^−1^ can be computed easily. The exception is where the joint distribution for *Z* is not known in closed form, but instead represented by samples from an MCMC simulation. This topic is taken up in Section 2.4.

When using QMC for EVPPI, we only apply it to the outer samples *X*, since in practice we do not need a great number of inner samples *Y* to achieve acceptable accuracy, that is of order of *ε*^−1^ samples as explained in Section 2.1. Simulating larger numbers of inner samples won’t be of benefit as it would only give a very accurate approximation for each fixed outer sample, and the variance of the outer sample continues to have a larger impact on the total error.

For QMC to achieve an MSE *ε*^2^, we still need the number of inner samples per outer sample to be an order of magnitude of *ε*^−1^, but only need around *ε*^−1^ outer samples, due to the properties of low-discrepancy sequences [26]. Hence the total computational cost is an order of around *ε*^−2^, compared to *ε*^−3^ for standard Monte Carlo. This is the around same cost as MLMC, as we will show in the context of two real applications in Section 3. As low-discrepancy sequences reduce variance in non-nested Monte Carlo, QMC reduces the computational cost to estimate the total EVPI. In our numerical tests we will include an assessment of this property.

However, similar to the standard MC method, QMC also fails to give an estimate of bias. In the numerical tests of this paper, we only use MLMC to estimate the bias and determine the number of inner samples required to bound the bias.

### 2.4 MLMC and QMC when parameters are sampled using Markov Chain Monte Carlo

As discussed in the introduction, cost-effectiveness decision models often rely on Bayesian evidence synthesis to inform parameters so that the joint distribution for some parameters may be represented by samples from an MCMC simulation. Model inputs based on MCMC samples cause three problems for MLMC and QMC estimation. Firstly, if the parameters estimated by MCMC are highly correlated then the number of MCMC samples needed for reliable inference may be large, due to inefficiencies with the MCMC sampler. We address this issue for both MLMC and QMC in Section 2.4.1. Secondly, because MCMC only provides samples and not a closed form representation of the joint distribution, we cannot form the inverse distribution which is needed for QMC. We address this issue Section in 2.4.1. Thirdly, because MCMC does not provide a closed form posterior distribution, we cannot sample from conditional distributions, which are needed for EVPPI estimation whether performed by standard nested MC, MLMC, or QMC. We address this by using multivariate approximations to the posterior distribution is described in Section 2.4.2.

#### 2.4.1 Random selection of MCMC samples for MLMC and QMC

To reduce the computational expense of generating MCMC samples, we first generate a sufficiently large number *N*_*c*_ of MCMC samples in advance to store in a data set, and randomly select samples from it. Our motivation is that we can then use uniformly distributed random variables for sampling, rather than needing further expensive MCMC. For MLMC, when constructing the level *𝓁* bias estimator 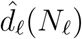 we generate 2^*𝓁*^ uniform random samples *U* ^(*n*)^ in [0, 1] and choose corresponding MCMC samples with index ⌊ *N*_*c*_*U* ^(*n*)^ ⌋ (the largest integer smaller than or equal to *N*_*c*_*U* ^(*n*)^). This ensure the same samples in the level *𝓁* and *𝓁* − 1 estimators.

For QMC, the idea is to sort the stored MCMC samples first, and QMC points denote the corresponding position in the sorted sequence. This has the added advantage that it removes the need for an inverse of the posterior distribution. For high dimensional problems, we use an efficient approach to sample a uniform distribution on a large empirical dataset. Given a large enough MCMC sample, we further select the samples we will use in the calculation from the MCMC sample pool. We apply QMC on the random selection. For details on how exactly to apply QMC on random selection, please refer to [32]. This algorithm uses samples from the Uniform distribution to repeatedly bisect the MCMC sample space until only a single sample is found (technical name is recursive spectral bisection algorithm for graph partitioning [33]). The order and elements of X on which to sort is determined by the first two dimensions from principal components analysis (PCA). Using only the first two elements was found to be sufficient for the applications in Section 3, and captured the majority of the impact of the variation of our parameters on the net benefit, and thus the impact on EVPPI. However, if many elements of X had high weights, indicating that the PCA cannot identify two significant dimensions, this approach would be not suitable.

#### 2.4.2 Multivariate approximation of MCMC to obtain conditional distributions

For EVPPI estimation, we need to sample from the distribution of *Y* conditionally on a specific value of *X*^(*n*)^. However this is challenging when *X* and *Y* are correlated and their joint distribution is only known through a MCMC sample. In theory, we simply need to set the value of *X*^(*n*)^ to be fixed and rerun the MCMC simulation to get *Y* ^(*n,m*)^. However, this is highly inefficient, since a separate burn-in is required for each individual sample of *X*^(*n*)^.

To solve this problem, we compute the mean *µ* and covariance matrix Σ of the MCMC samples *Z*, and use the multivariate normal distribution (MVN) 𝒩(*µ*, Σ) to approximate the posterior distribution. Then the distribution of *Y* ^(*n,m*)^ conditional on *X*^(*n*)^ is known to be another MVN [34]. The benefit is that the random variable we need to generate is Normally distributed and we can apply our MLMC and QMC formulations.

Note that, the MVN approximation is only suitable for parameters whose joint posterior is a symmetric unbounded distribution. Parameters such as hazard ratios or odds ratios will need to be log transformed to achieve this, as we do in the examples in the following sections. Heavy tailed distributions may be better represented by some other closed form multivariate distribution, such as multivariate t-distribution. However, our approach to MCMC will only work if a transformation to some closed form multivariate distribution (with closed form conditional distributions) is possible.

## 3 Applications

### 3.1 Simplified cost-effectiveness model in depression

We applied our MLMC and QMC EVPPI estimators to an artificial model comparing options for the treatment of depression. We adopted the decision tree structure illustrated in Figure 3. This is based on a previously published model but is populated with artificial quality of life and costs of outcomes as well as a Bayesian NMA, implemented using MCMC, of response and relapse outcomes from constructed randomised controlled trial (RCT) data [35]. There are three options compared: no treatment, Cognitive Behavioural Therapy (CBT), and antidepressants. We label these options *d* = 1, 2, 3 respectively. The same structure is used for both treated (by any treatment) and untreated patients but the probabilities of recovery and relapse depend on the treatment. In this model, patients begin their treatment (or no treatment) in the “Depressed” node and move to “Recovery” if they have an initial response to treatment and “No recovery” if not. Following “Recovery”, patients can either experience a relapse and end in the “Relapse” node or remain healthy in the “No relapse” node.

**Figure 3:**
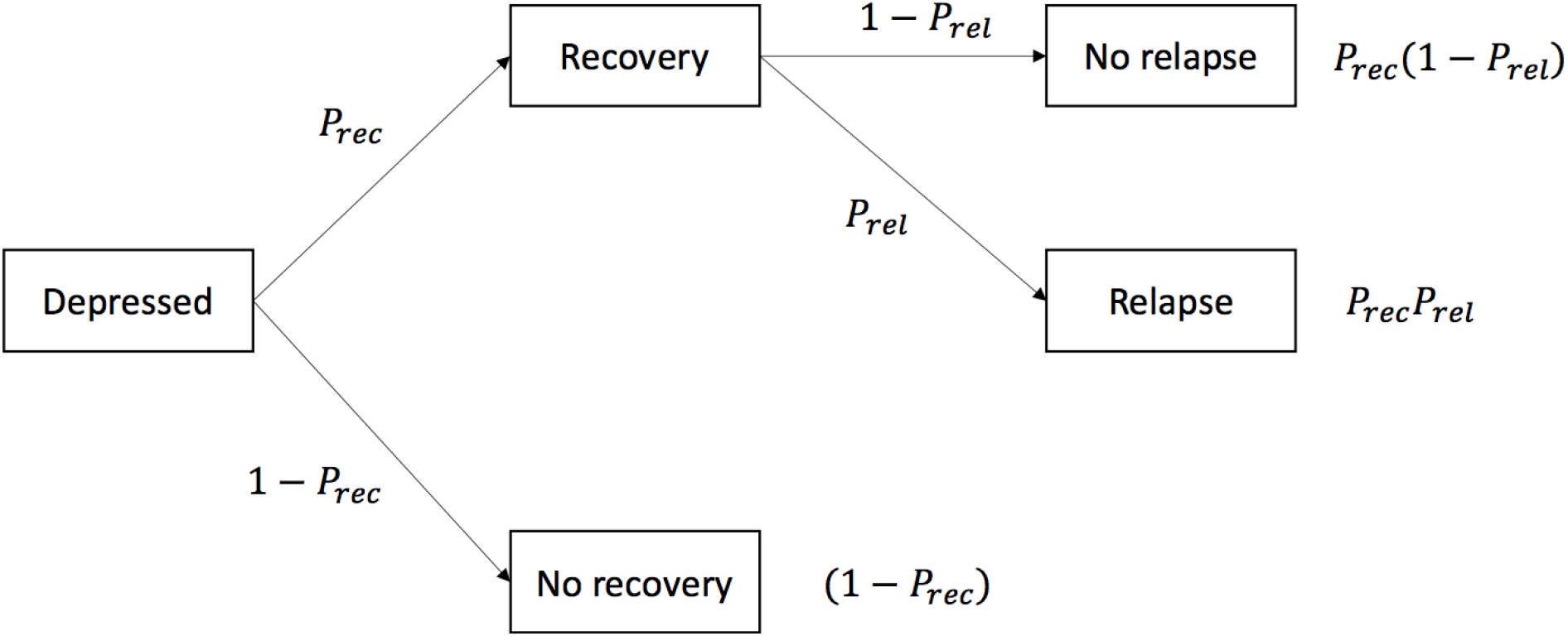
Decision tree for depression toy model (The probabilities are defined in Table 2).

Costs and Quality Adjusted Life Years (QALYs) associated with 30-years (approximately lifetime) in the final states are assumed to follow normal distributions (Table 1).

**Table 1:**
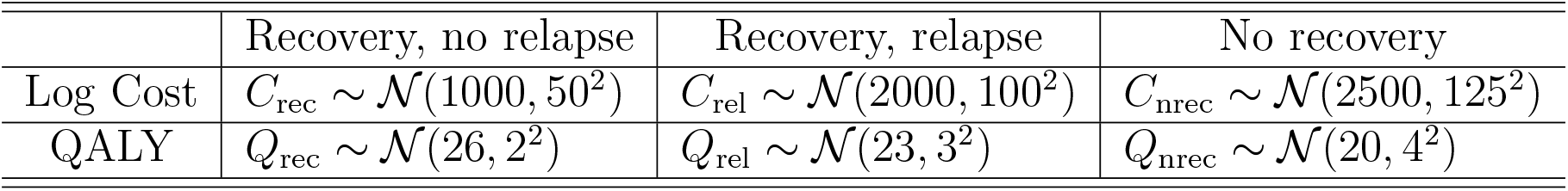
30-year costs and QALYs for depression toy model.

**Table 2:** Probabilities of events for depression toy model. The expit is the inverse of the logistic link function with definition 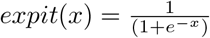. “lor” is the log odds ratio.

| $d$ | Treatment | $P_{\text{rec}}^{(d)}$ | $P_{\text{rel}}^{(d)}$ |
| --- | --- | --- | --- |
| 1 | No treatment | Beta(6, 200) | Beta(2, 100) |
| 2 | CBT | $\text{expit}(\text{logit}(P_{\text{rec}}^{(1)}) + \text{lor}_{\text{rec}}^{(2)})$ | $\text{expit}(\text{logit}(P_{\text{rel}}^{(1)}) + \text{lor}_{\text{rel}}^{(2)})$ |
| 3 | Antidepressant | $\text{expit}(\text{logit}(P_{\text{rec}}^{(1)}) + \text{lor}_{\text{rec}}^{(3)})$ | $\text{expit}(\text{logit}(P_{\text{rel}}^{(1)}) + \text{lor}_{\text{rel}}^{(3)})$ |

As given in Table 2, we assumed Beta distributions for the probabilities of relapse and recovery on no treatment. Log odds ratios of recovery and relapse on CBT and antidepressants come from two NMAs, each of which consist of 5 trials based on constructed data. These data were analysed using a Bayesian binomial outcomes logistic link NMA implemented in the OpenBUGS software version 3.2.3 rev 1012 [19, 36], which generated MCMC samples of the posterior distributions for the log odds of relapse and recovery. As conditional distributions are necessary for EVPPI, we used the multivariate Normal distributions discussed in Section 3.3.2 to produce the following approximate posterior distributions.

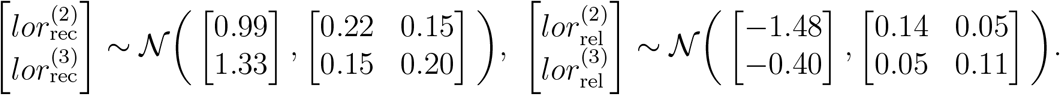

The net benefit for treatment option *d* for a patient in our model is

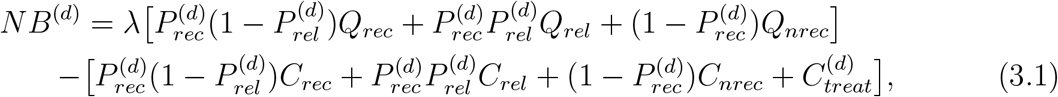

where *C*_*treat*_ = (0, 300, 30) is the fixed initial treatment cost.

In our case, *f*_*d*_(*Z*) = *NB*^(*d*)^, the net benefit of treatment *d* given

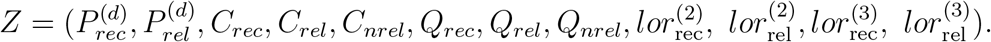

We estimate the EVPPI for four subsets *X* of *Z*. These are the probabilities of recovery and relapse 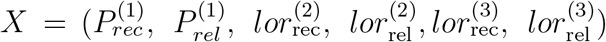, the costs and QALYs *X* = (*C*_*rec*_, *C*_*rel*_, *C*_*nrel*_, *Q*_*rec*_, *Q*_*rel*_, *Q*_*nrel*_), the log odds ratios for CBT 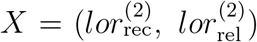, and the log odds ratios for anti-depressants 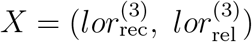.

### 3.2 Real, complex health economic model of DOACs in atrial fibrillation

In the previous decision tree example, the NB function was simple and quick to evaluate. In this section we present an example where the net benefit function is more complex, so that generating a sample of NBs is more computationally expensive. Thus we might expect that standard nested MC methods require unrealistic sample sizes for a required accuracy. The MCMC sampler required more simulations to characterise the posterior distribution, since a statistical model of higher dimension was used, and the sampled chains were more strongly autocorrelated. This example is a model recently used to compare the cost-effectiveness of DOACs for prevention of stroke in atrial fibrillation, the full details of which have been published [18, 17]. In brief, this was a Markov model with 17 health states, including death, a 3-month cycle length, lifetime horizon, and taking the UK National Health Service perspective. It compared first-line treatment by coumarin with international normalised ratio 2-3 to four DOACs: apixaban 5mg twice daily (BD), dabigatran 150mg BD, rivaroxaban 20mg once daily (OD), and edoxaban 60mg OD in 70 year old adults. Treatment switching from DOAC to coumarin or no treatment or from coumarin to no treatment was modelled. This model included four events with long-term impacts on costs, QALYs, and event risks: ischemic stroke, myocardial infarction (MI), major extracranial bleed, and intracranial haemorrhage (ICH). Health states were defined by which of these states a patient had experienced, giving the model structure illustrated in Figure 4. In addition, transient ischemic attack (TIA) and systemic embolism (SE) were modelled as transient events with no long-term consequences. Relative effects were informed by a systematic literature review of RCTs comparing DOACs and coumarin. Log hazard ratios relative to coumarin on the seven possible events were estimated using a Bayesian competing risks NMA conducted in OpenBUGS. Log hazard ratios for no treatment compared to coumarin came from a NMA of previously identified RCTs [37]. Log hazards of events on coumarin were estimated using a Bayesian natural history model fit to the coumarin arms of the RCTs [38], thus providing absolute probabilities of events on all possible treatments. MCMC samples from the posterior distributions of these three Bayesian analyses were input directly to the cost-effectiveness model. Costs, utilities, and impact of event history on current risks were estimated using closed form distributions fit to data identified in targeted literature reviews. Full details of all parameters and model assumptions are provided in previous publications [18, 17].

**Figure 4:**
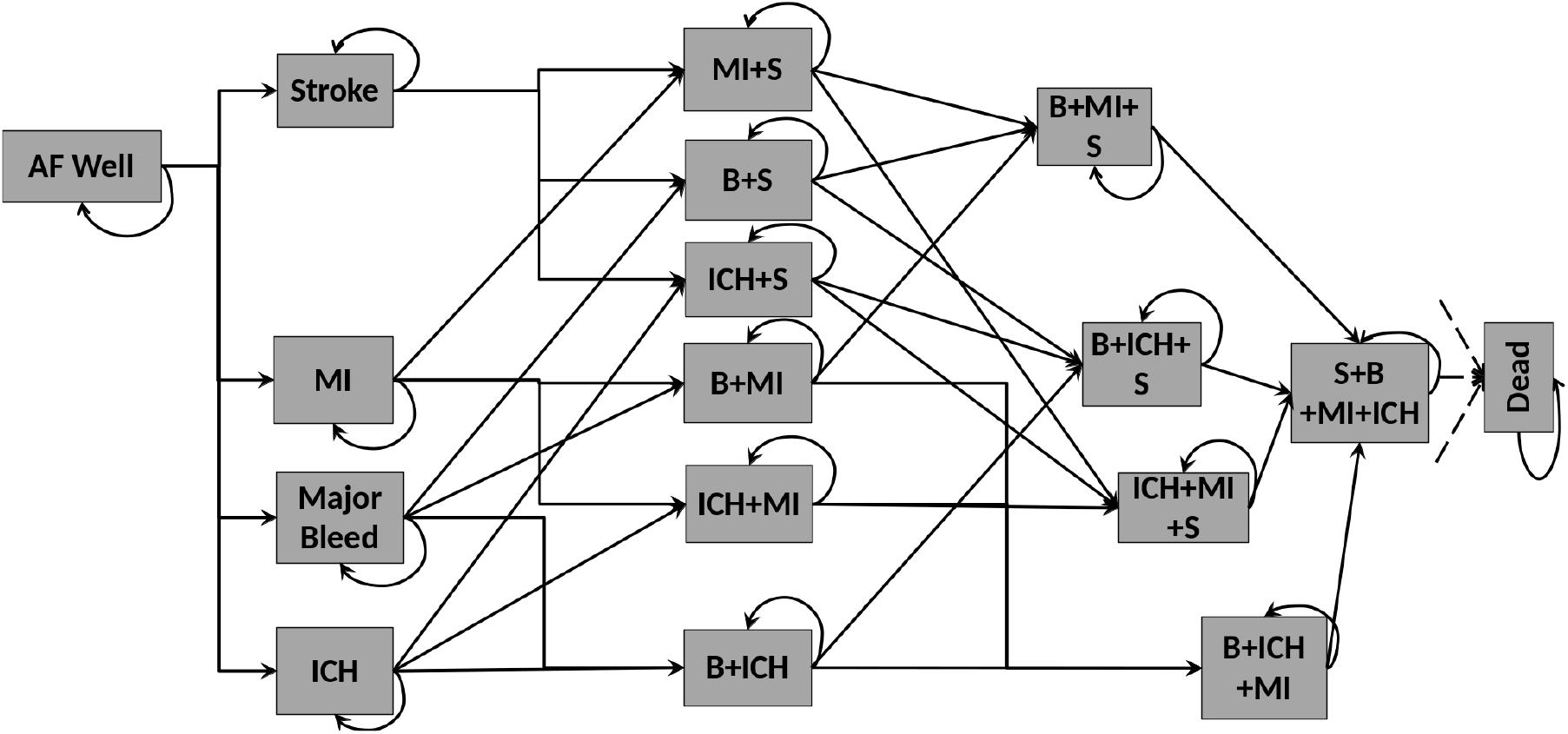
Structure of the DOACs for prevention of stroke in atrial fibrillation cost-effectiveness model. AF=Atrial fibrillation, MI=Myocardial infarction, ICH=Intracranial haemorrhage, B=Major bleed, S=Stroke

In addition to the EVPPI for subsets corresponding to categories of the model parameters (e.g. those generated by MCMC, or treatment switching probabilities), we would like to investigate the potential value of conducing a trial comparing apixaban and dabigatran (the two best DOACs) with a coumarin control arm. We consider two such designs. First is a simple trial that records only the log hazard ratios of apixaban and dabigatran compared to coumarin. Second is a complex trial that records these hazard ratios but also gathers evidence on the absolute probabilities on coumarin, all event and management costs, all event disutilities, all state utilities, and the treatment switching probabilities.

## 4 Results

### 4.1 MLMC and QMC results for simplified cost-effectiveness model in depression

For comparison, we set the MSE to be 0.25 (*ε* = 0.5) by bounding the bias by 0.25 and variance by 0.1875. Table 3 shows the number of samples needed as the computational cost to achieve a 0.25 MSE.

**Table 3:** Comparison of computational cost measured in units of 10^6^ samples. *P*_*rec*_ = probability of recovery, *P*_*rel*_ = probability of relapse following recovery, *lor*_*rec*_ = log odds ratios of recovery for CBT and antidepressants compared to no treatment, *lor*_*rel*_ = log odds ratios of relapse for CBT and antidepressants compared to no treatment, MC=Standard nested Monte Carlo, MLMC=Multilevel Monte Carlo, QMC=Quasi Monte Carlo

| EVPPI | Estimates | 95% Credible interval | Computational Cost |  |  |
| --- | --- | --- | --- | --- | --- |
|  |  |  | MC | MLMC | QMC |
| $P_{rec}^{(d)}, P_{rel}^{(d)}, lor_{rec}^{(2)}, lor_{rel}^{(2)}$<br>$lor_{rec}^{(3)}, lor_{rel}^{(3)}$ | 275 | [274, 277] | 15.14 | 26.93 | 5.63 |
| Cost and QALYs | 287 | [286, 289] | 13.02 | 22.85 | 5.12 |
| $lor_{rec}$ and $lor_{rel}$ CBT | 7 | [6, 9] | 787.20 | 109.90 | 8.19 |
| $lor_{rec}$ and $lor_{rel}$ anti-depressants | 1 | [0, 3] | 78.90 | 61.86 | 6.23 |

From Table 3, we can see that QMC achieves the same degree of accuracy with lower computational cost than standard nested MC and MLMC when the optimal number of inner samples is determined, since QMC has smallest number in Computational Cost Columns in the table. MLMC only starts to show computational savings relative to standard nested MC for the calculation of EVPPI for *lor*_2_ and *lor*_3_, since these parameters are correlated. Further cost-effectiveness results are provided in Appendix 7.2.1.

In order to reproduce the complexity that may be encountered in a real cost-effectiveness model, we extended our example from 3 to 20 treatment options. The model structure, outcome costs, and outcome QALYs remained the same but treatment effects and costs were randomly generated. Both MLMC and QMC continue to work well and QMC still has lowest computational costs given the number of inner samples. Full details of the model and EVPPI results are provided in Appendix 7.2.2 and 7.2.3.

### 4.2 Comparison with regression approximation methods on simplified cost-effectiveness model in depression

We also calculated the EVPPI using the regression approaches of generalised additive models (GAM) and Gaussian processes (GP), as implemented in the R code of Sheffield Accelerated Value of Information (SAVI), and stochastic partial differential equations integrated nested laplace approximations (INLA-GP), as implemented in the R package BCEA [12, 14, 39]. Since the GP method requires large matrices to be inverted, SAVI is restricted to the first 7500 samples from an economic model for this method, while BCEA struggles with larger samples due to memory restrictions; also, the primary advantage of these methods is their ability to estimate EVPPI with fewer samples and thus lower computational cost. For these reasons, we used only 7500 samples for the comparison. To provide an approximately fair comparison, we restricted MC, QMC and MLMC to 50,000 samples, which has a similar computation time to running 7500 samples followed by GAM, GP, and INLA-GP regression. Note that SAVI automatically uses GP for parameter sets larger than 5 and in this example, with 6 parameters GAM methods are computationally infeasible [13], while the BCEA implementation of GAM also suffers with memory restrictions when applied to larger parameter sets. As a consequence, only the GP-based methods are possible for estimating the EVPPI for the two sets of six parameters. The R code of SAVI provides the estimates of both standard error (SE), i.e. the standard deviation of the estimator, and the upward bias [13]. Similar to MLMC and QMC, the MSE of SAVI would be the sum of the variance of the estimator and the square of the upward bias. Furthermore, BCEA INLA-GP cannot estimate standard errors.

Table (4) suggests that both GP and GAM perform better for 2-dimensional sets of log odds ratios, giving estimates close to the reference and small standard errors. On the more complex 6-dimensional set of probabilities and the 6-dimensional cost and QALYs, QMC and MLMC are close to the reference MC estimate but only QMC offers computational savings (lower RMSE) over standard MC. On these 6-dimensional sets, the point estimate for probabilities of GP is in agreement with those of MC methods, although the RMSE are larger and estimates appear to be biased. The point estimate for cost and QALYs of GP has a much smaller bias and standard error, but is not consistent with the reference MC estimate. INLA-GP agrees on the 6-dimensional probabilities but gives a poor estimate of the EVPPI of the costs and QALYs. This was indicated by the BCEA diagnostic quantile-quantile plots (qq-plots), which suggested poor fit of the underlying regression model for INLA-GP (details in Appendix 7.3).

**Table 4:** Comparison of estimates and uncertainties for EVPPIs in depression toy model. Use 50,000 samples for MC, MLMC and QMC, and 7500 samples for GAM, GP and INLA-GP. MC reference value is that from Table 3.

| Parameters<br>(size) | MC<br>reference<br>(RMSE)<br>(Bias, SE) | MC<br>(RMSE)<br>(Bias, SE) | QMC<br>(RMSE)<br>(Bias, SE) | MLMC<br>(RMSE)<br>(Bias, SE) | GAM<br>(RMSE)<br>(Bias, SE) | GP<br>(RMSE)<br>(Bias, SE) | INLA-GP |
| --- | --- | --- | --- | --- | --- | --- | --- |
| Probabilities<br>(6) | 275<br>(0.5)<br>(0.25, 0.43) | 273.62<br>(5.19)<br>(2.59, 4.48) | 281.20<br>(3.03)<br>(2.59, 1.57) | 274.84<br>(12.00)<br>(6.00, 10.38) | NA | 322.19<br>(100.17)<br>(89.66, 44.68) | 293.44 |
| Costs & QALYs<br>(6) | 287<br>(0.5)<br>(0.25, 0.43) | 286.86<br>(5.46)<br>(2.73, 4.72) | 286.93<br>(4.58)<br>(2.73, 3.68) | 285.13<br>(9.80)<br>(4.90, 8.48) | NA | 557.03<br>(0.77)<br>(0.09, 0.76) | 547.42 |
| CBT<br>(2) | 7<br>(0.5)<br>(0.25, 0.43) | 29.53<br>(20.47)<br>(9.72, 18.01) | 44.65<br>(20.44)<br>(9.72, 17.98) | 22.03<br>(19.44)<br>(9.72, 16.82) | 12.26<br>(10.52)<br>(5.52, 8.96) | 11.91<br>(12.26)<br>(1.74, 12.14) | 13.83 |
| Antidepressant<br>(2) | 1<br>(0.5)<br>(0.25, 0.43) | 0.28<br>(18.73)<br>(8.00, 16.94) | 5.12<br>(16.58)<br>(8.00, 14.52) | 4.10<br>(16.00)<br>(8.00, 13.84) | 1.56<br>(18.10)<br>(14.20, 11.23) | 6.62<br>(10.95)<br>(4.87, 9.81) | 4.81 |

The EVPI and its RMSE for the depression model were 574.91 and 3.12 using standard MC, respectively, and 575.47 and 2.37 using QMC. The lower RMSE suggests computational savings from using QMC.

### 4.3 MLMC and QMC results for real, complex health economic model in atrial fibrillation

In summary, the model parameters include 32 where uncertainty is represented by normal random variables, 8 by uniform random variables, 15 by beta distributed random variables, and 42 by 3 sets of MCMC samples (7-dimensional MCMC samples for baseline log hazard ratios, 28-dimensional MCMC samples for log hazard ratios relative to the baseline, and 7-dimensional MCMC samples for log hazard ratios of no treatment).

Table 5 shows the EVPPI estimates computed using the standard MC method, and the relative computational cost required by MLMC and QMC to produce estimates with the same degree of accuracy. MLMC is much more efficient than standard MC when EVPPI is large. In cases where small sample exploration indicated a small or zero EVPPI, we only use standard MC. QMC also works better than standard MC for all the EVPPIs.

**Table 5:** EVPPI of the DOACs model: comparison of computational cost (measured in units of 10^5^ samples). QMC couldn’t give an estimate with *ε*^2^ MSE within a reasonable time due to the large number of inverse normal and beta function evaluations. MLMC couldn’t provide estimates for EVPPI in reasonable time when values are nearly 0.

| EVPPI | Value | $M$ | RMSE | Bias | Sd | Computational Cost ($10^5$ samples) | | |
| --- | --- | --- | --- | --- | --- | --- | --- | --- |
|  |  |  |  |  |  | MC | MLMC | QMC |
| Simple Trial | 196 | 2048 | 1.23 | 0.25 | 1.20 | 2144 | 68.63 | – |
| Complex Trial | 273 | 256 | 1.23 | 0.25 | 1.20 | 157 | 33.78 | 89.54 |
| All cost | 0 | 256 | 0.91 | 0.91 | 0 | 2.62 | – | 2.62 |
| Switch probability | 0 | 128 | 0.01 | 0.01 | 0 | 1.31 | – | 1.31 |
| All MCMC | 348 | 256 | 1.23 | 0.25 | 1.20 | 139.9 | 24.47 | 31.72 |
| Baseline MCMC | 0 | 128 | 0.79 | 0.57 | 0.54 | 1.31 | – | 0.97 |
| Loghr MCMC | 286 | 256 | 1.23 | 0.25 | 1.20 | 317.1 | 31.85 | 21.21 |
| No treatment MCMC | 0 | 128 | 1.05 | 1.05 | 0 | 1.31 | – | 1.31 |
| All Utility | 0 | 256 | 0.70 | 0.70 | 0 | 2.62 | – | 2.62 |

Comparing MLMC and QMC, we observe that MLMC performs better for the cases where the parameters are correlated (Simple & Complex Trials), and the QMC works relatively better for the other cases.

### 4.4 Comparison with regression approximation methods on real, complex health economic model in atrial fibrillation

As in section 4.2, we compared our estimates with those provided by GP and INLA-GP, as implemented in SAVI and BCEA, respectively. We conducted comparisons on the large parameter sets where MLMC was necessary. In this case GAM could not be applied as there are too many parameters to identify a regression model without making excessively strong assumptions (such as additivity and linearity) about the relationship between the input parameters and the conditional expected net benefit. Again to compare the relative accuracy of the different methods given the same computational cost, we restricted MC, QMC and MLMC to 50,000 samples and GP and INLA-GP to 7500. Results are presented in Table 6.

**Table 6:** Comparison of estimates and uncertainties for EVPPIs in DOACs model. MC reference value is from Table 5, using sufficient samples to achieve pre-specified RMSE. Use 50,000 samples for MC, MLMC and QMC and 7500 samples for GP and INLA-GP. MC reference value is that from Table 5.

| Parameters<br>(size) | MC<br>reference<br>(RMSE)<br>(Bias, SE) | MC<br>(RMSE)<br>(Bias, SE) | QMC<br>(RMSE)<br>(Bias, SE) | MLMC<br>(RMSE)<br>(Bias, SE) | GP<br>(RMSE)<br>(Bias, SE) | INLA-GP |
| --- | --- | --- | --- | --- | --- | --- |
| Simple Trial<br>(14) | 196<br>(1.23)<br>(0.25, 1.20) | 201.93<br>(52.70)<br>(11.63, 51.40) | 192.02<br>(17.14)<br>(11.63, 12.59) | 194.80<br>(23.25)<br>(11.63, 20.12) | 406.72<br>(417.07)<br>(412.29, 62.95) | 741.09 |
| Complex Trial<br>(51) | 273<br>(1.23)<br>(0.25, 1.20) | 248.37<br>(38.91)<br>(14.52, 36.10) | 251.73<br>(32.40)<br>(14.52, 28.96) | 259.40<br>(29.04)<br>(14.52, 25.12) | 742.10<br>(327.23)<br>(323.21, 51.14) | - |
| All MCMC<br>(40) | 348<br>(1.23)<br>(0.25, 1.20) | 354.22<br>(13.68)<br>(4.94, 12.76) | 361.21<br>(11.13)<br>(4.94, 9.97) | 367.31<br>(9.87)<br>(4.94, 8.55) | 642.95<br>(140.05)<br>(130.88, 49.83) | 256.56 |
| Loghr MCMC<br>(28) | 286<br>(1.23)<br>(0.25, 1.20) | 264.49<br>(16.13)<br>(7.84, 14.10) | 266.68<br>(14.63)<br>(7.84, 12.35) | 291.12<br>(15.68)<br>(7.84, 13.56) | 944.69<br>(522.37)<br>(517.42, 71.76) | 551.98 |

This application was more challenging for all methods. The INLA-GP could not estimate for the complex trial. The GP results were severely biased upwards compared to MC, probably due to the poor high-dimensional regression approximation. In addition, they lack face validity as the parameters included in the Loghr MCMC case are a subset of those in the all MCMC case, so the loghr MCMC case should not have a higher EVPPI. The INLA-GP estimates also disagreed with the reference results for all methods, with the possible exception of the MCMC set. However, a poor fit of the underlying regression model was only identified by diagnostic plots in the All MCMC case. The MLMC and QMC methods using only 50,000 samples gave estimates close to the MC reference value, and had RMSEs lower than standard MC.

Again testing testing QMC for estimating total EVPI, we found the EVPI and standard deviation were 416.79 and 5.38 using standard MC and 418.12 and 2.41 using QMC. As in the depression example, this represented reduced bias and variance.

## 5 Discussion

This paper has developed more efficient Monte Carlo sampling methods to estimate EVPPI in complex and realistic cost-effectiveness models. We have generalised the previously published MLMC estimator for EVPPI to models in which the distributions of input parameters are only known through MCMC samples and demonstrated that it can be much more efficient than standard MC for computing many-parameter EVPPI in a realistically complex economic model. We have also provided the first implementation of QMC to EVPPI estimation. Unlike previous work on efficient EVPPI estimation via model regression and INLA-GP [12, 39, 13], we have separately quantified both bias and variance of our estimators and included them in our credible intervals. We have compared the accuracy of the QMC and MLMC methods relative to the GAM, GP, and INLA-GP regression approaches for the same approximate computational cost. The MLMC estimator can easily give an estimate of the bias and this can be used to obtain credible intervals for MLMC but also for both QMC and standard nested MC estimators, so long as MLMC is conducted in addition to either QMC or MC.

The main contributions of this paper have been to extend MLMC for EVPPI and develop QMC for EVPPI. While our results suggest that QMC and MLMC can provide substantial computational savings over MC, and greater accuracy than regression techniques, we have explored only two example models. A more formal investigation would be needed to fully compare MC, MLMC, QMC, and regression. This could involve building a range of models with increasing numbers of health states, input parameters and decision options, and explore higher correlation and greater MCMC on the input parameters. Further theoretical research is also required to understand why each of the methods performs well under different circumstances. Without such a program, it would be inappropriate to generalise and make firm recommendations. However, we make some observations based on our empirical findings and the theoretical understanding of QMC and MLMC.

Our two examples may indicate a general trend that QMC outperforms MLMC when the underlying model is simple (e.g. decision tree or Markov model with few states, few parameters, low correlation) and MLMC outperforms QMC when the model is complex (e.g. Markov model with many states, large numbers of correlated or MCMC parameters). Our depression toy example also indicated that MC can outperform MLMC when the models are simple. If an estimate of the bias is required, MLMC must be employed as MC, QMC, and regression cannot estimate the bias. Furthermore, if very high accuracy, or low bias, are required MLMC is likely best as higher levels of MLMC will eventually achieve any accuracy; however, this may not be computationally feasible. Conversely, if the EVPPI is very small, which could be found by an initial run of MC with few samples, then MLMC may offer limited computational savings over MC. Indeed, we found in the depression toy example that MLMC could not provide an estimate for small EVPPI in a reasonable time. Theoretically, QMC should be no worse than MC in all cases. However, the computational saving depends on the specific case. In practice, furthermore, QMC can perform worse than MC, as was seen for the simple trial example in the DOACs model where our implementation failed to produce an estimate in a reasonable time as too many inverse distribution function evaluations were needed.

We have also found that MLMC and QMC provide more reliable and computationally efficient estimates of the EVPPI than regression techniques when parameter sets are large and when the EVPPI value is large and a precise EVPPI estimate is required. Conversely, and in line with MLMC theory, we did not find a huge advantage over standard MC or approximation methods when the EVPPI or parameter sets are small. We also do not expect advantage of MLMC or QMC when applied to single parameter EVPPI. However, we found using QMC conferred computational savings over standard MC when estimating the total EVPI.

From our experiment, our MLMC and QMC methods do not rely on many approximations or distributional assumptions, to incorporate inputs whose distribution is estimated by MCMC. Note that in QMC we use PCA to identify which of the first two dimensions of *X* to sort on. The reason we use two dimensions is that we find for this problem the first two dimensions of PCA gives an reasonably good approximation and it is not computationally expensive. Still, it was necessary to assume a MVN approximation. This was required for sampling from conditional distributions where parameters were correlated in both MLMC and QMC, and to estimate the inverse cumulative distributions for QMC. We presented a method of resampling from the MCMC samples using random or quasi-random numbers, thus avoiding the need for large numbers of MCMC samples or the inverse cumulative distribution, although this does not address the need for correlated samples. The MVN approximation is likely suitable for parameters such as relative treatment effects expressed as log odds or hazard ratios [36]. As an alternative, we explored multivariate t-distributions to capture possible fat tails [40]. However, the credible interval largely coincided with what was provided by the MVN approximation, see Appendix 7.5 for detailed results.

A primary disadvantage of the MLMC method from an applied perspective is the requirement for more than a single random sample from a standard probabilistic sensitivity analysis. The model regression approaches of GP, GAM, or INLA-GP require only samples from the input parameters and the estimated costs and QALYs to provide estimates of EVPPI. MLMC requires implementation of a more complicated form of nested simulation than in standard Monte Carlo, plus sampling from conditional distributions to provide the necessary estimates. This challenge may limit its applicability. We have provided R code in the appendix for both the depression and DOACs model, which users can adapt to their own models. QMC also requires conditional sampling but can use the same code as standard nested Monte Carlo; it requires only a switch to quasi-random numbers for the random number generators (example code also provided in the appendix). However, we found QMC not to work as well in the case of highly complex cost-effectiveness models such as Markov models.

Although we have used very large numbers of simulations (10^5^ to 10^8^ samples), for trial funding decisions, such large numbers of samples and low RMSE may not be necessary. Such decisions are made by applying the population EVPI and EVPPI to the cost of a proposed trial. The population EVPI and EVPPI are the per-person values we have discussed but scaled to the annual number of patient who would benefit scaled and discounted over the technology lifetime for which we expect the research to be relevant. In the DOACs example, if we assume 5000 patients per year, discounting at 1.035, and summing over a technology lifetime of 10 years gives a total population of approximately 43038 patients. Scaling the value of a “Simple trial” comparing apixaban to dabigatran estimated by only 50,000 MC samples from Table 6 gives a population EVPPI of £8.69 million and RMSE of £2.27 million. This implies lower and upper 95% credible bounds of about £4.25 million and £13.14 million, respectively. If an RCT comparing these DOACs cost £3 million, it would be below the lower bound and the decision would be to fund; no greater accuracy or lower variance would be needed. However, trial costs and EVPPI may be closer in other situations.

Our work so far has been limited to EVPPI, but to truly determine whether a future study will be cost-effective we would need an estimate of the EVSI [5]. Although the EVPPI for a set of parameters may be large, the EVSI for all but impractical study designs could be small. Despite its importance, EVSI is rarely estimated due to unfamiliarity with the skills required, and, for trials potentially informing many parameters, high computational requirements [41]. Efficient sampling schemes, such as importance sampling, Gaussian approximation, and moment matching approaches have been explored, but a general solution applicable to all model and trial complexities remains elusive [42, 43, 11]. Implementing QMC for EVSI would be straightforward, although the computational savings are unknown. Conversely, constructing an MLMC estimator, necessary for bias estimation, would require considerable research effort. However, MLMC and QMC may provide an accurate and efficient estimator of EVSI and improve its adoption by the heath economic community.

## 6 Conclusion

In this paper, we developed MLMC and QMC to the computation of EVPPIs and applied to a decision tree and Markov model example. In some cases, both methods improved the computational efficiency of the standard nested MC method, although they are more difficult to implement than standard MC. We found that for small numbers of parameters and small EVPPI values, GAM and GP were sufficient for EVPPI estimation. However, for large numbers of parameters and EVPPI values, where GAM is not feasible, MLMC and QMC can give substantially more accurate and precise estimates than GP and INLAGP. Further theoretical and empirical research is required to make formal recommendations between standard nested MC, QMC, MLMC, and the regression techniques.

## Data Availability

All code and data necessary to the produce the results are available in this GitHub repository: https://github.com/Bogdasayen/Example_MLMC_and_QMC_for_EVPPI
The simple demo of MLMC can be found at this repository: https://github.com/Bogdasayen/mlmc_evppi_demo

https://github.com/Bogdasayen/mlmc_evppi_demo

## Acknowledgement

WF, ZW, and HT were supported by the Hubs for Trials Methodology Research (HTMR) network grant N79 for this work. HT and NJW were supported by the HTMR Collaboration and innovation in Difficult and Complex randomised controlled Trials In Invasive procedures (ConDuCT-II). HT and NJW were also supported by the National Institute for Health Research (NIHR) Bristol Biomedical Research Centre (BRC) for part of this work. HT was furthermore supported by MRC grant MR/S036709/1. CA would like to thank support of EPSRC EP/R018561/1 Bayes4Health. CJ was funded by the UK Medical Research Council programme MC UU 00002/11. The directly acting oral anticoagulants for prevention of stroke in atrial fibrillation model was funded by NIHR Health Technology Assessment programme project number 11/92/17 and NIHR Senior Investigator award NF-SI-0611-10168.

We are also grateful to Mark Strong at University of Sheffield for providing his R code to estimate EVPPI, plus its SE and upward bias, using GAM and GP.

## 7 Appendix

### 7.1 Further details of MLMC

#### 7.1.1 Estimating number of levels *L* and samples *N*_*l*_ required for MLMC

As mentioned, the number of levels *L*, and the number of outer samples *N*_*𝓁*_ in each level, can be tuned so that the MLMC estimator achieves a specific balance of bias, precision and computational speed. For example, we might want to minimise the cost to obtain a RMSE of at most *ε* and a bias of at most *ε/*2.

It was shown in [23] that the expectation and variance of the level *𝓁* estimators are approximately *C*_1_2^−*α𝓁*^ and *C*_2_2^−*β 𝓁*^. Therefore we can estimate log(*C*_1_) and −*α* as the intercept and slope of a linear regression fitted to the three data points from levels *𝓁* = 0, 1, 2 with log *d*_*𝓁*_ as the outcome and *𝓁* as the predictor, similarly for log(*C*_2_) and −*β*. It was shown in [23] for the standard MLMC method (Section 2.2) we have *α* = 1 due to the central limit theorm, *β* = 1 due to the smoothness of the net benefit function *f*_*d*_. Alternative sampling schemes where *α* and *β* are not 1 are possible, and discussed in later sections, and in these cases the procedure below can be used to estimate them by regression.

The step-by-step procedure is as follows:

1. Run a small pilot sample, for example of size *N*_*l*_ = 1000 for each *𝓁* and estimate the expectation and variance at the first three levels *𝓁* = 0,1,2 (i.e. 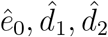). At *𝓁* = 1, for example, the expectation would be 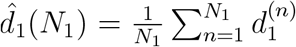 and the variance 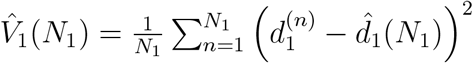.
2. Use regression on the three points to estimate *C*_1_ (and *α* if not equal to 1) so that the level *𝓁* expectation of bias reduction is approximately *C*_1_2^−*α𝓁*^
3. Use regression on the three points to estimate *C*_2_ (and *β* if not equal to 1) so that the level *𝓁* variance is approximately *C*_2_2^−*β𝓁*^. The variance of the MLMC estimator is therefore 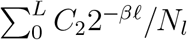
4. Choose level *L* large enough to control the bias of the final EVPPI estimator, which is *C*_1_2^−*αL*^ ≤ *ε/*2
5. Choose *N*_*𝓁*_ for each level *𝓁* to minimise the total computational cost 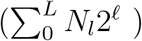 while achieving RMSE *ε*, which requires knowledge of the constants *C*_2_ and *β*. This minimisation can be done by constrained optimisation or by trial and error.

The details of these calculations are given in [20, 21] and illustrated in the supplementary code in section 7.4 of this paper.

Two additional refinements of the MLMC method are explained in the appendix 7.1.2 and 7.1.3. The first is a method of arranging the random samples of *X* and *Y* to ensure a higher correlation between successive estimators *ê*_*𝓁*−1_(*N*), *ê*_*𝓁*_(*N*) for which it was shown *β* = 1.5 [21, 44]. The second is an alternative method of constructing the series for MLMC estimator, and is preferable when the EVPPI is large compared with the EVPI.

#### 7.1.2 Further details of the antithetic variable approach to the level estimator

In this appendix we discuss how to enable 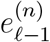 and 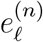 have a higher correlation by utilising all the inner samples. For 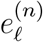 we need to generate 2^*𝓁*^ samples of *Y* ^(*n,m*)^ conditional on *X*^(*n*)^. That is

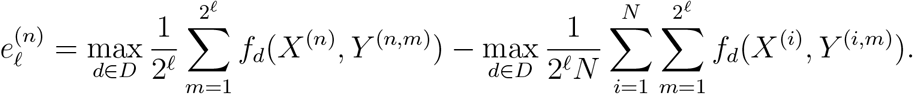

For 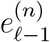, we use the same *X*^(*n*)^ and separate the 2^*𝓁*^ samples of *Y* ^(*n,m*)^ into two sets, both of which have 2*𝓁*−1 samples, to construct two samples and get the average for the first term and use all the samples for the second term. That is

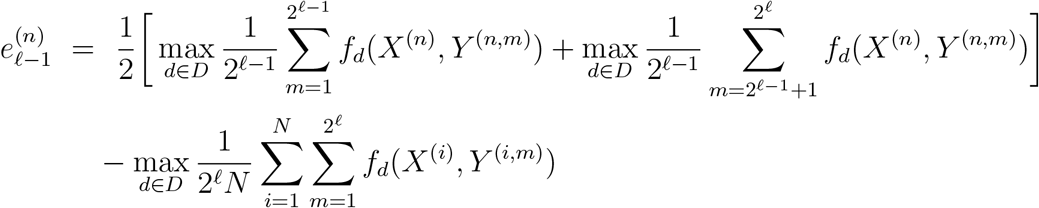

Then the difference estimator 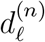 becomes

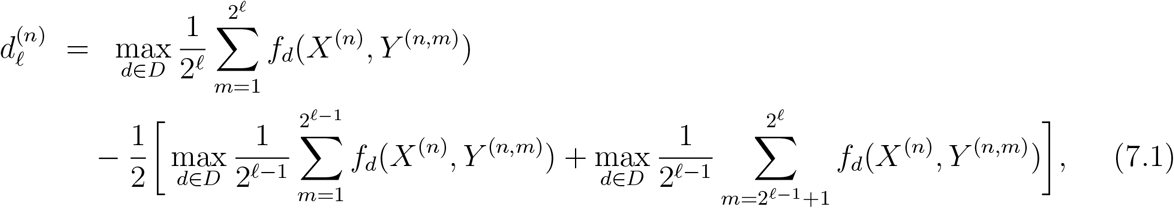

Note now that the second term of 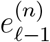 and 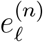 cancels. If all three terms have the same optimal choice *d*^*^, then 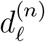 becomes 0. Theorem 3 in [23] shows that the variance of the new 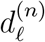 is the order of *O*(2^−3*𝓁/*2^).

#### 7.1.3 MLMC for DIFF

As illustrated in Figures 5(a) and 5(b), it is possible for the EVPPI to be large or small compared with EVPI, respectively. If The EVPI value is known, and it is known that the EVPPI is close in value to the EVPI, it will require fewer samples to estimate DIFF=EVPIEVPPI as its smaller magnitude usually leads to a smaller variance. This appendix describes how to construct an estimator for DIFF.

**Figure 5:**
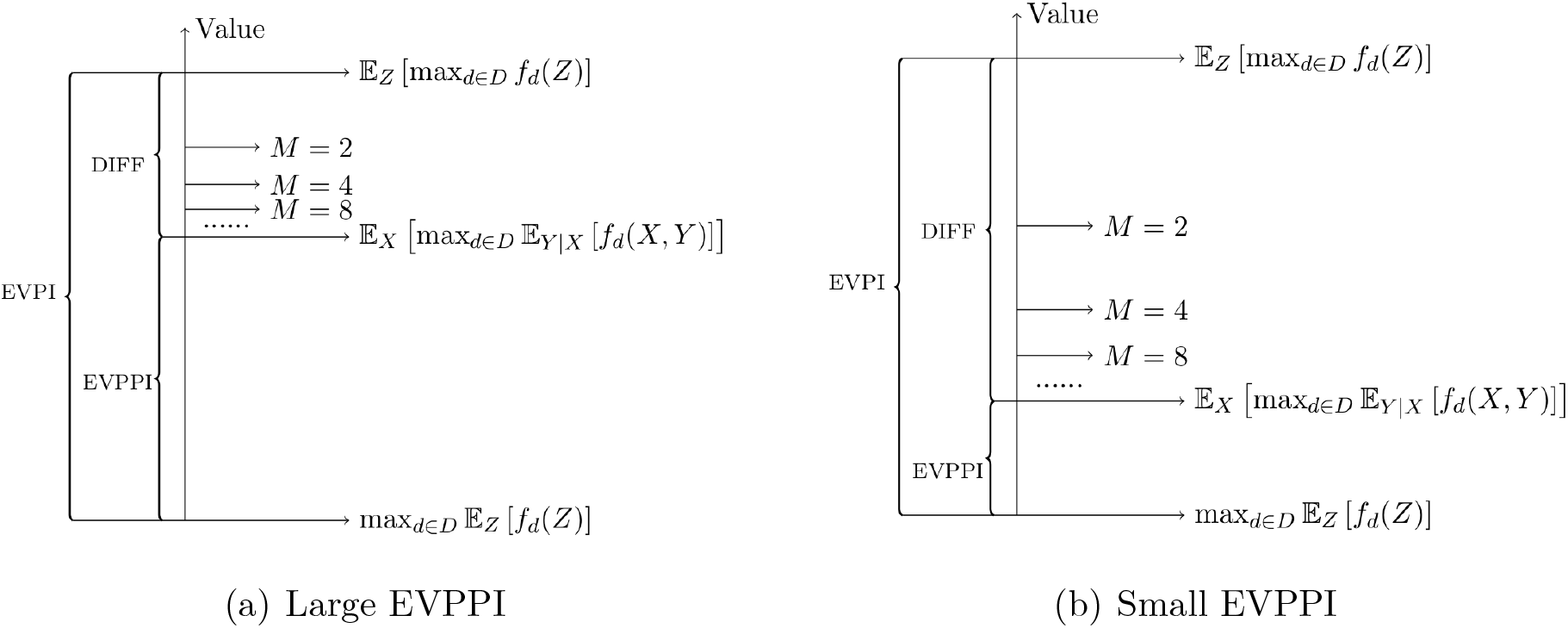
Structure of large or small EVPI

**Figure 6:**
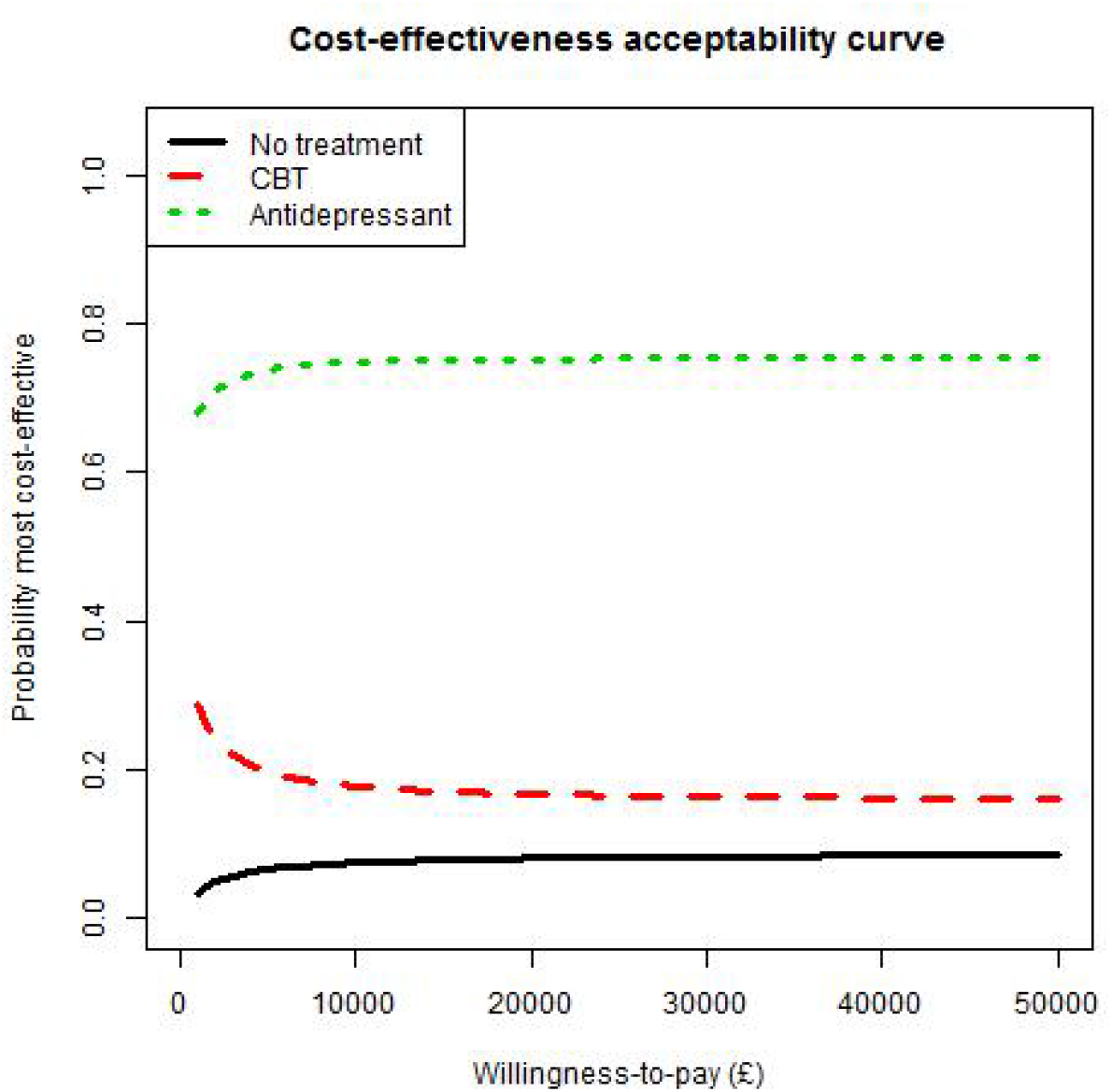
Cost-effectiveness acceptability curve. Each line represents the probability that the corresponding treatment has highest net benefit at each willingness-to-pay threshold.

By definition, we have

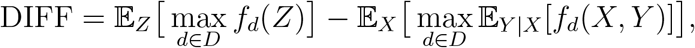

and the standard nested MC estimator is

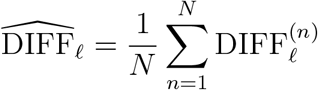

Where

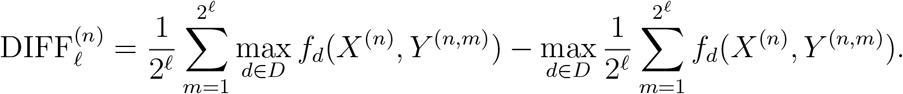

For MLMC, we find that 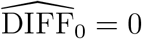 and the corresponding MLMC estimator of DIFF is

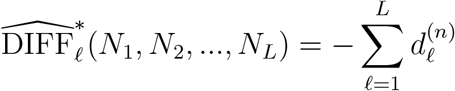

where 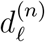 is the same as (7.1) since the first term of the 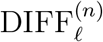 and 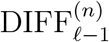 are cancelled out similar to the previous subsection. Compared with (2.7), we get rid of *ê*_0_(*N*_0_). Therefore, from a practical point of view, it is always easier to estimate a smaller quantity since the variance would be lower. For larger EVPPI as shown in Figure 5(a), it is more reasonable to estimate DIFF instead of estimating EVPPI directly.

### 7.2 Further details of the depression cost-effectiveness toy model

#### 7.2.1 Cost-effectiveness results from the depression toy model

CBT and antidepressants are cheaper than no treatment but antidepressants are the cheapest. CBT and antidepressants confer greater benefits (QALYs) but antidepressants are most effective. Incremental net benefit is highest with antidepressants at a willingness-to-pay threshold (*λ*) of £20,000. The CEAC indicates that antidepressants have a high probability (*>* 75%) of being most cost-effective at all realistic willingness-to-pay thresholds.

#### 7.2.2 Details of the 20 treatment option depression model

In order to explore the complexity that may be encountered in a real cost-effectiveness model, we extended our example from 3 to 20 treatment options. The model structure, outcome costs, and outcome QALYs for each treatment remained the same but treatment effects and costs were randomly generated. However, log odds of relapse and recovery were sampled from a Normal distribution with mean 0 and standard deviation 1 and drug costs were either *£* 500, *£* 1000, *£* 2000, or *£* 3000 with simple rules ensuring treatments with high recovery and low relapse and most expensive, and vice versa (Table 8). The NMA was also extended to 50 trials of up to 5 arms comparing a random selection of the 20 treatments; baseline log odds response and recovery were sampled from Normal with mean 0 and standard deviation 0.25, while numbers recovering or relapsing were sampled from binomials with the appropriate probabilities (Figure 7(a) and Figure 7(b) present network plots for this constructed data).

**Table 7:** Cost-effectiveness results of the 3-treatment depression toy model.

|  | No treatment | CBT | Antidepressant |
| --- | --- | --- | --- |
| Total costs (£) | 2566.79<br>(2252.17, 2934.14) | 2490.75<br>(2148.05, 2874.31) | 2452.64<br>(2093.18, 2844.76) |
| Total QALYs | 20.16<br>(12.53, 27.76) | 20.47<br>(13.16, 27.68) | 20.62<br>(13.49, 27.64) |
| Incremental costs (£) | 0<br>(0, 0) | -76.04<br>(-443.73, 287.29) | -114.15<br>(-537.83, 239.71) |
| Incremental QALYs | 0<br>(0, 0) | 0.30<br>(-0.12, 1.31) | 0.46<br>(-0.19, 1.80) |
| Net Benefit<br>($\lambda$=£20,000) | 400694<br>(247991.10, 552672.40) | 406863.20<br>(260751.50, 551144.90) | 409983.90<br>(267368.00, 550330.30) |
| Incremental NB<br>($\lambda$=£20,000) | 0<br>(0, 0) | 6169.19<br>(-2342.05, 26456.00) | 9289.87<br>(-3672.06, 36367.09) |

**Table 8:** Treatment effects and costs of the 20-treatment depression model.

|  | Relapse LOR | Recovery LOR | Price (£) |
| --- | --- | --- | --- |
| Treatment 1 | 0.00 | 0.00 | 500 |
| Treatment 2 | -0.16 | 0.10 | 1000 |
| Treatment 3 | 0.93 | 1.37 | 2000 |
| Treatment 4 | 0.59 | 0.84 | 1000 |
| Treatment 5 | 0.28 | 0.58 | 1000 |
| Treatment 6 | 0.26 | -1.97 | 500 |
| Treatment 7 | -0.01 | -0.60 | 1000 |
| Treatment 8 | 0.72 | 1.92 | 2000 |
| Treatment 9 | -0.44 | -0.26 | 1000 |
| Treatment 10 | 0.81 | 1.85 | 2000 |
| Treatment 11 | -1.43 | -0.89 | 2000 |
| Treatment 12 | -0.41 | 0.56 | 1000 |
| Treatment 13 | -0.93 | 0.12 | 1000 |
| Treatment 14 | -0.49 | 0.45 | 1000 |
| Treatment 15 | -1.65 | 1.15 | 3000 |
| Treatment 16 | -0.36 | 0.29 | 1000 |
| Treatment 17 | 0.40 | -0.22 | 500 |
| Treatment 18 | 0.58 | -0.46 | 500 |
| Treatment 19 | 0.13 | 0.75 | 1000 |
| Treatment 20 | 0.49 | -2.08 | 500 |

**Figure 7:**
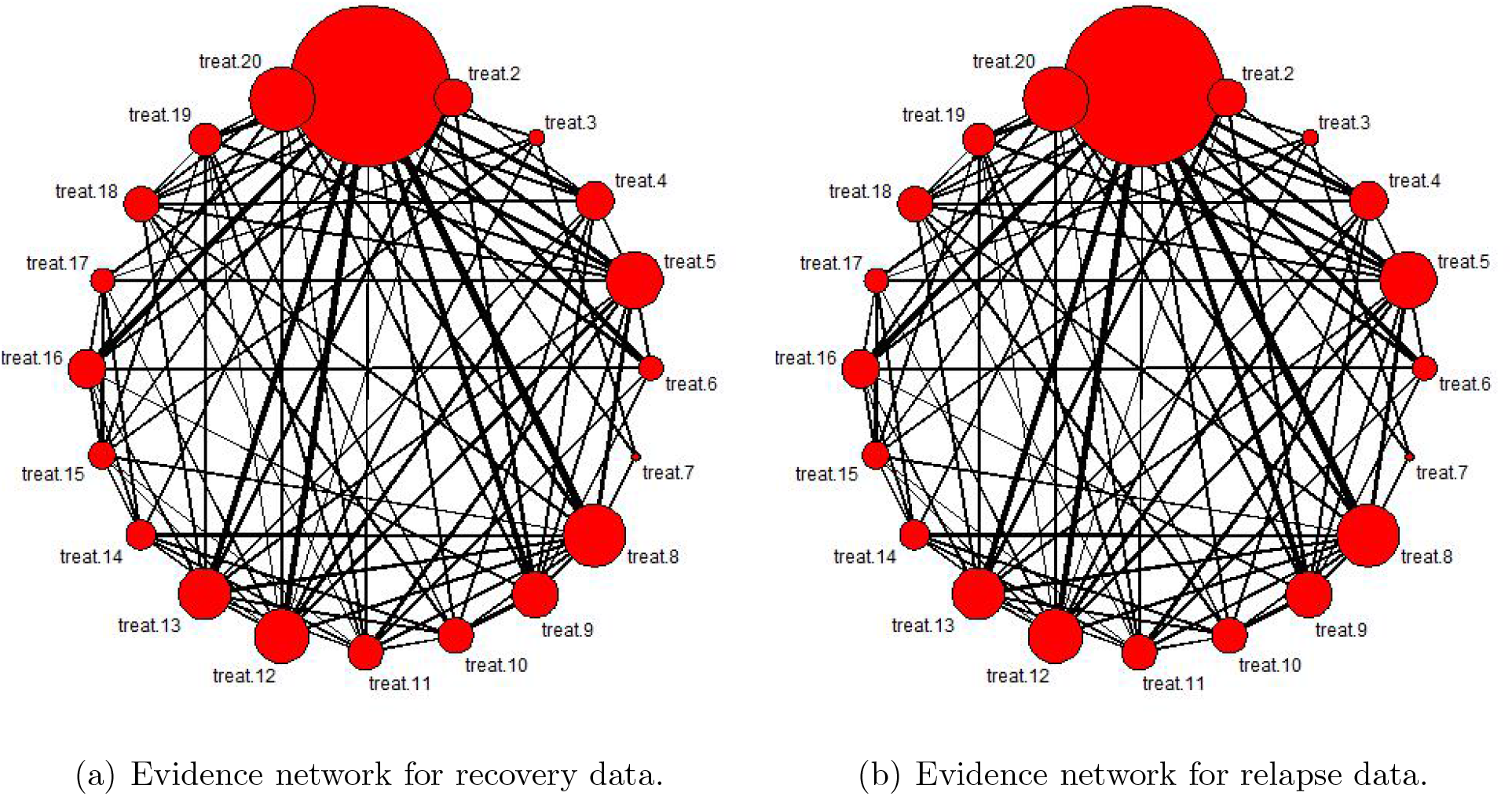
Evidence networks for recovery and relapse data in 20 treatment depression model. Each node represents a treatment being compared. An edge represents a trial comparing the treatments represented by the two connected nodes. If a path can be found between two nodes, the corresponding treatments can be compared by network meta-analysis.

The cost-effectiveness results are presented in Table 9, with CEACs presented in Figure 8. Treatment 10, which had a high log odds ratio of both recovery (1.85) and relapse (0.81) and a high cost (£2000), had the highest expected incremental net benefit (*£*14590.49 with 95% Credible Interval (−8706.228, 46266.03)) and greatest probability (*>* 60%) of being most cost-effective at all willingness-to-pay thresholds.

**Table 9:** Cost-effectiveness results from the 20-treatment depression toy example.

|  | Total costs | Total QALYs | Incremental costs | Incremental QALYs | Net Benefit | Incremental NB |
| --- | --- | --- | --- | --- | --- | --- |
| Treatment 1 | 2956.657<br>(2716.0, 3197.1) | 20.22<br>(12.7, 27.7) | 0<br>(0, 0) | 0<br>(0, 0) | 401453.1<br>(252603, 551490.9) | 0<br>(0, 0) |
| Treatment 2 | 3449.322<br>(3208.0, 3689.9) | 20.24<br>(12.8, 27.7) | 492.66<br>(482.1, 498.8) | 0.029<br>(-0.012, 0.107) | 401541.1<br>(253699.6, 550615.2) | 88.0<br>(-746.9, 1672.0) |
| Treatment 3 | 4342.823<br>(4094.0, 4592.8) | 20.67<br>(13.7, 27.5) | 1386.16<br>(1288.5, 1455.1) | 0.452<br>(-0.215, 1.38) | 409123.7<br>(271047.1, 547659.8) | 7670.6<br>(-5692.4, 26406.3) |
| Treatment 4 | 3394.855<br>(3150.8, 3638.7) | 20.46<br>(13.2, 27.6) | 438.19<br>(382.86, 476.54) | 0.245<br>(-0.114, 0.753) | 405924.7<br>(262204.9, 549397.4) | 4471.6<br>(-2719.8, 14672.2) |
| Treatment 5 | 3422.529<br>(3181.2, 3662.5) | 20.35<br>(13.0, 27.6) | 465.87<br>(433.91, 487.43) | 0.135<br>(-0.0610, 0.4232) | 403691<br>(257802.8, 550271.7) | 2237.9<br>(-1680.1, 8042.3) |
| Treatment 6 | 2994.332<br>(2750.6, 3237.4) | 20.07<br>(12.4, 27.7) | 37.67<br>(13.8, 74.4) | -0.149<br>(-0.469, 0.0673) | 398434.7<br>(245141.4, 552521.3) | -3018.3<br>(-9446.1, 1303.0) |
| Treatment 7 | 3477.776<br>(3236.4, 3719.0) | 20.13<br>(12.5, 27.7) | 521.11<br>(507.6, 541.9) | -0.0834<br>(-0.262, 0.037) | 399263.4<br>(247884.1, 551555.2) | -2189.6<br>(-5770.9, 235.8) |
| Treatment 8 | 4270.117<br>(4008.8, 4531.8) | 20.96<br>(14.3, 27.5) | 1313.46<br>(1169.9, 1423.1) | 0.740<br>(-0.349, 2.21) | 414947.7<br>(281912.9, 546828.5) | 13494.6<br>(-8262.5, 43019.6) |
| Treatment 9 | 3464.967<br>(3223.3, 3705.6) | 20.18<br>(12.6, 27.7) | 508.31<br>(502.7, 517.2) | -0.0329<br>(-0.107, 0.0153) | 400285.6<br>(250357.9, 551439.2) | -1167.4<br>(-2656.7, -203.8) |
| Treatment 10 | 4256.8<br>(3995.6, 4522.0) | 21.01<br>(14.4, 27.5) | 1300.14<br>(1149.1, 1417.0) | 0.794<br>(-0.371, 2.37) | 416043.5<br>(283951.5, 546673.7) | 14590.4<br>(-8706.2, 46266.0) |
| Treatment 11 | 4481.95<br>(4239.8, 4723.1) | 20.12<br>(12.5, 27.7) | 1525.29<br>(1509.3, 1549.7) | -0.100<br>(-0.316, 0.0452) | 397923.8<br>(245962.3, 550628.6) | -3529.2<br>(-7869.5, -618.2) |
| Treatment 12 | 3419.747<br>(3175.8, 3659.0) | 20.36<br>(13.0, 27.6) | 463.09<br>(427.6, 486.3) | 0.145<br>(-0.066, 0.456) | 403904.2<br>(258407.7, 550235.4) | 2451.1<br>(-1791.7, 8693.1) |
| Treatment 13 | 3454.433<br>(3214.4, 3694.1) | 20.22<br>(12.7, 27.7) | 497.77<br>(490.9, 502.5) | 0.00855<br>(-0.0132, 0.0490) | 401126.4<br>(252442.7, 550957.8) | -326.6<br>(-767.6, 487.4) |
| Treatment 14 | 3432.998<br>(3191.5, 3673.1) | 20.31<br>(12.9, 27.6) | 476.34<br>(453.1, 491.2) | 0.0932<br>(-0.0405, 0.295) | 402841<br>(256229.7, 550297.2) | 1387.8<br>(-1288.1, 5446.3) |
| Treatment 15 | 5361.133<br>(5112.2, 5608.3) | 20.59<br>(13.5, 27.6) | 2404.47<br>(2320.1, 2463.3) | 0.3760<br>(-0.163, 1.14) | 406570<br>(266626.8, 547233.4) | 5116.9<br>(-5666.6, 20639.7) |
| Treatment 16 | 3437.581<br>(3196.8, 3676.8) | 20.29<br>(12.9, 27.6) | 480.92<br>(461.0, 493.3) | 0.0750<br>(-0.0333, 0.241) | 402473.2<br>(255667.2, 550211.3) | 1020.1<br>(-1147.8, 4357.3) |
| Treatment 17 | 2963.146<br>(2722.1, 3203.6) | 20.19<br>(12.7, 27.7) | 6.48<br>(1.8, 14.2) | -0.0254<br>(-0.0858, 0.0109) | 400936.7<br>(251155.1, 551836.7) | -516.3<br>(-1726.7, 213.0) |
| Treatment 18 | 2972.343<br>(2731.7, 3212.8) | 20.15<br>(12.6, 27.7) | 15.6<br>(5.7, 31.1) | -0.0617<br>(-0.195, 0.0270) | 400201.6<br>(249608.4, 552002.6) | -1251.4<br>(-3941.0, 527.4) |
| Treatment 19 | 3411.843<br>(3168.7, 3651.9) | 20.39<br>(13.1, 27.6) | 455.18<br>(413.1, 483.1) | 0.177<br>(-0.0803, 0.551) | 404543.8<br>(259755.6, 549912.4) | 3090.7<br>(-2054.1, 10598.3) |
| Treatment 20 | 2994.37<br>(2750.6, 3237.6) | 20.07<br>(12.4, 27.7) | 37.71<br>(13.8, 74.2) | -0.149<br>(-0.469, 0.066) | 398432.8<br>(245143.9, 552534.5) | -3020.2<br>(-9461.8, 1287.2) |

**Figure 8:**
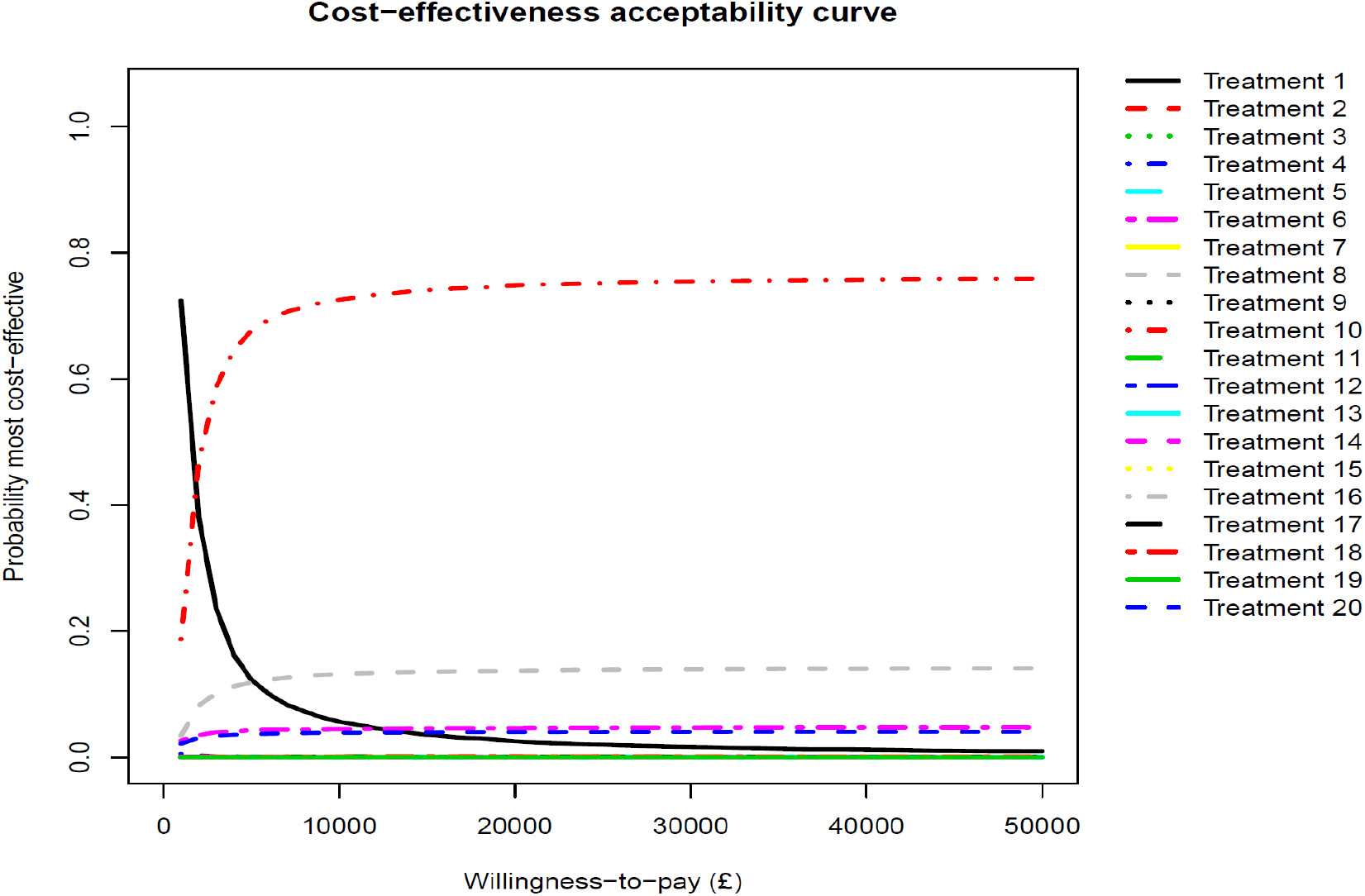
Cost-effectiveness acceptability curves (for any treatment with a probability of being most cost-effective greater than 5% at some threshold) for the 20-treatment depression model.

**Figure 9:**
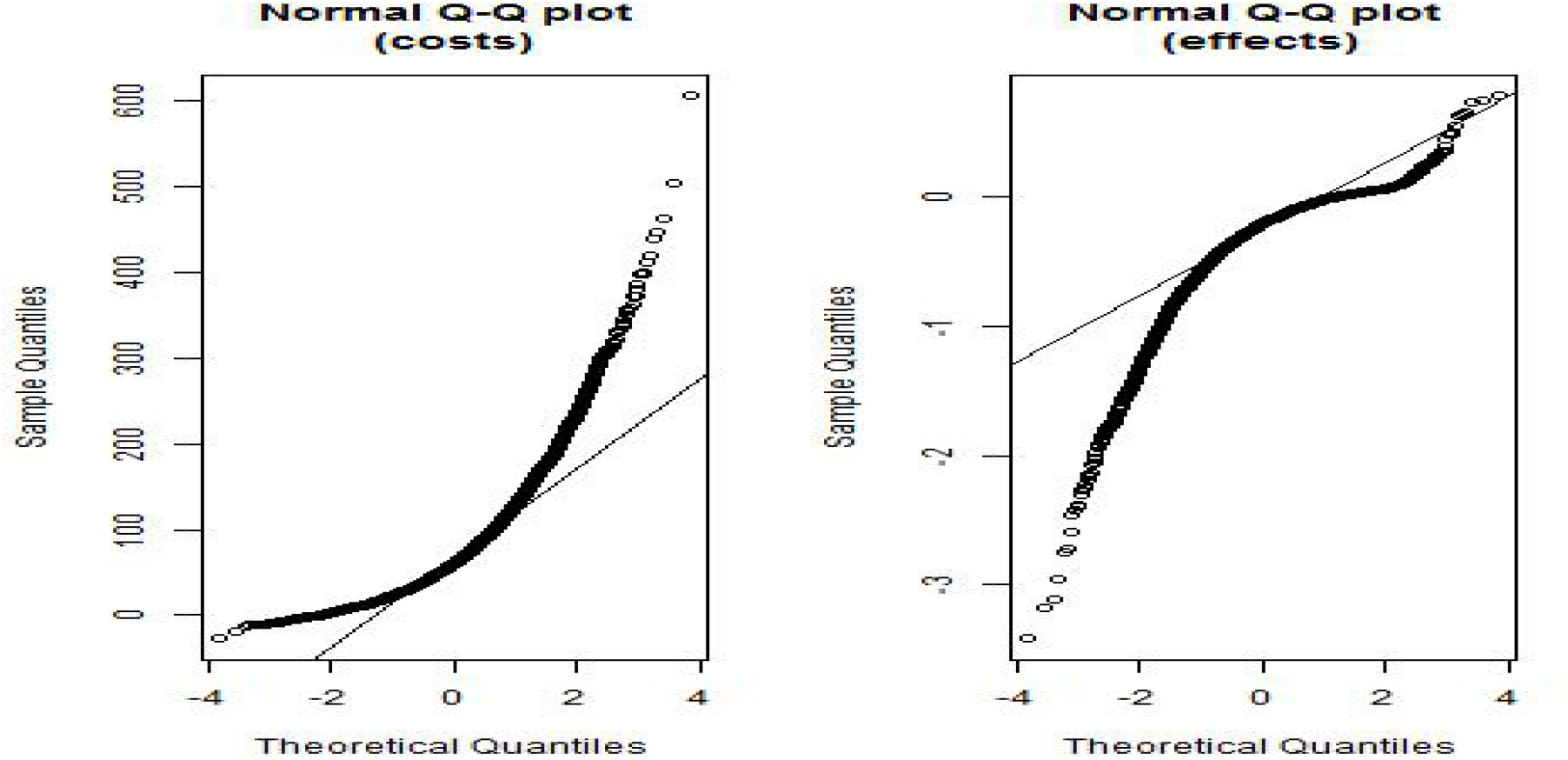
QQ-plots for INLA-GP estimate of EVPPI for costs and QALYs set in depression toy example

**Figure 10:**
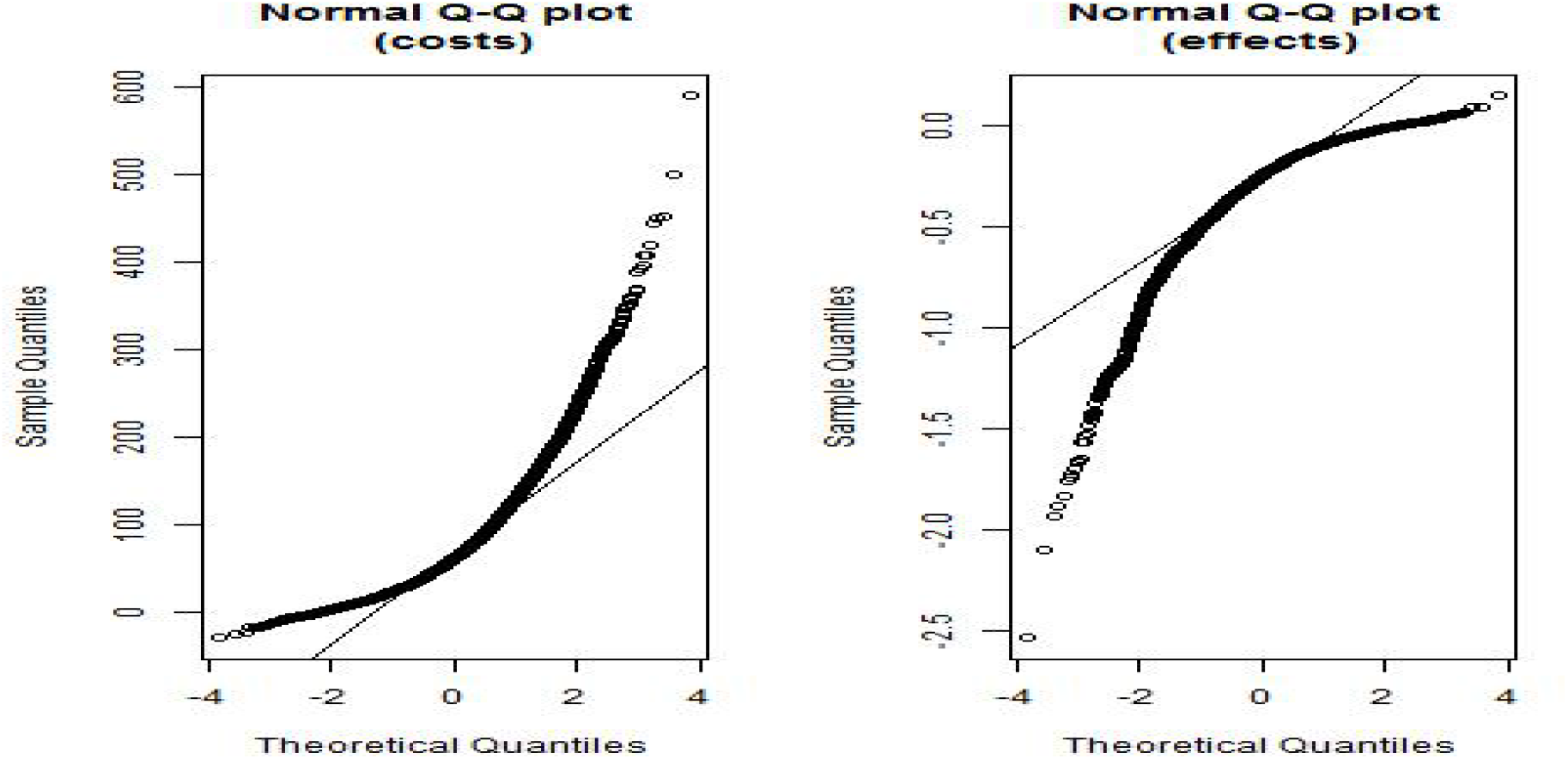
QQ-plots for INLA-GP estimate of EVPPI for probabilities of relapse and recovery in depression toy model

**Figure 11:**
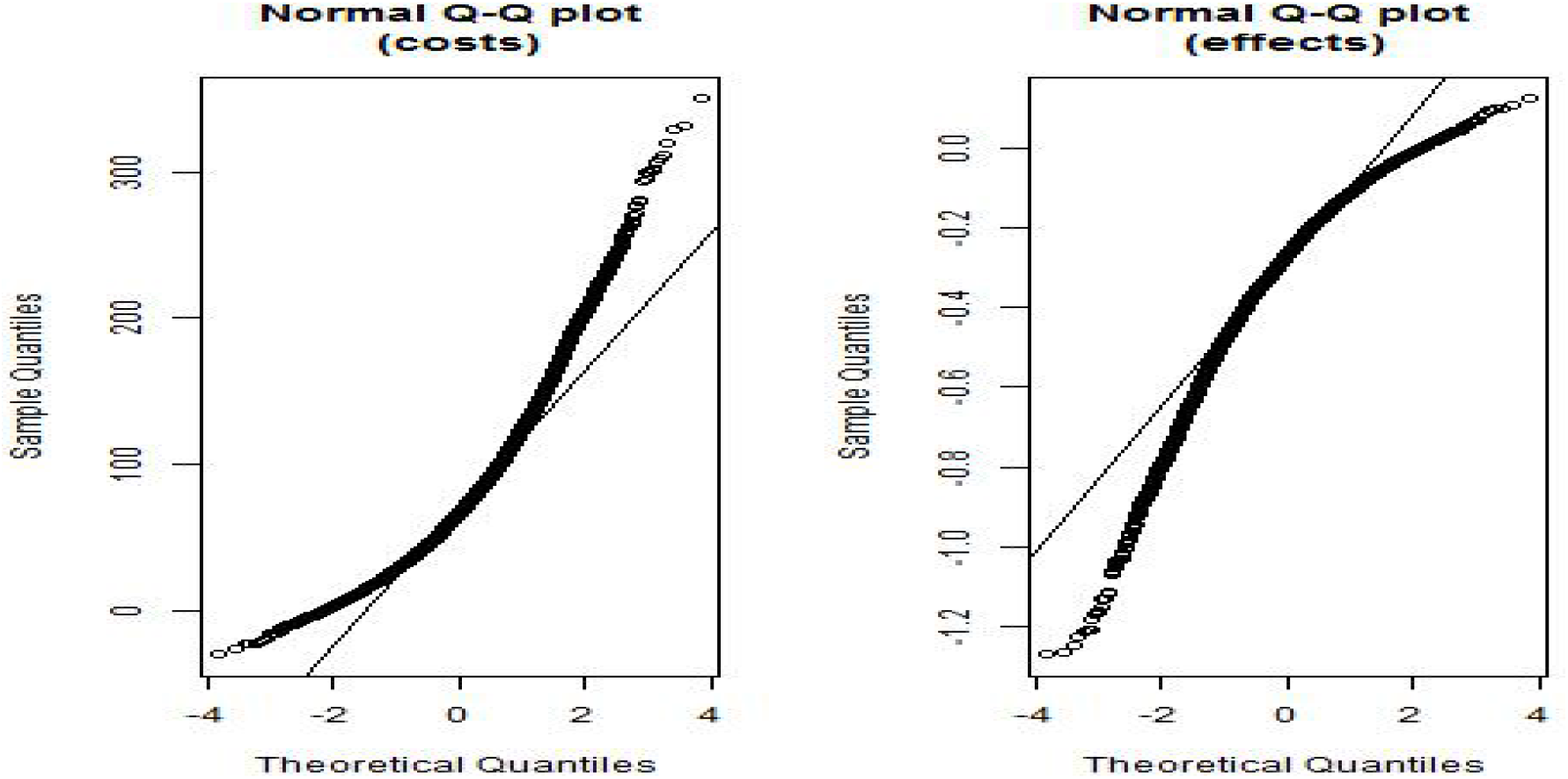
QQ-plots for INLA-GP estimate of EVPPI for CBT relapse and recovery log odds ratios in depression toy model

**Figure 12:**
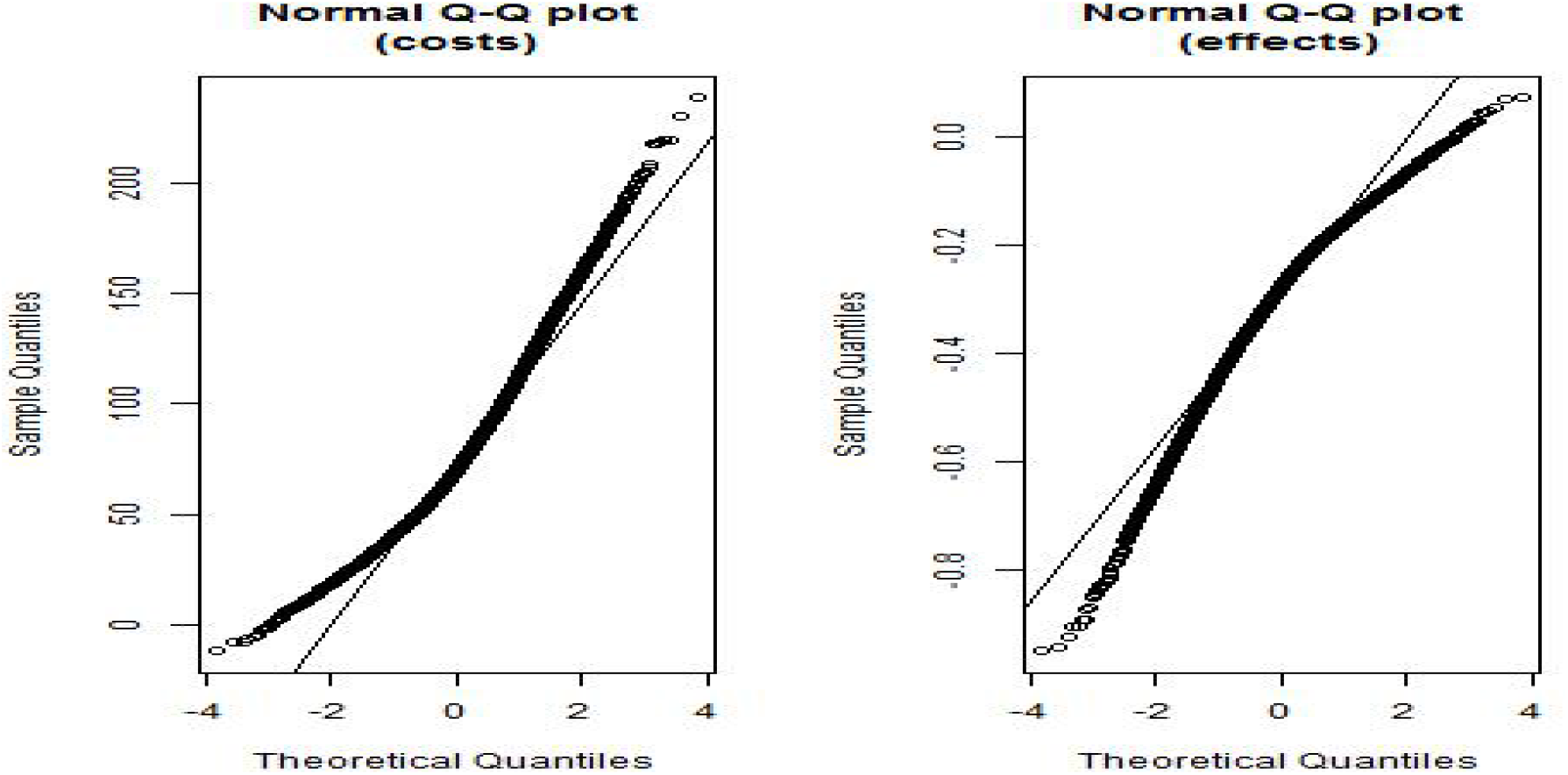
QQ-plots for INLA-GP estimate of EVPPI for antidepressant relapse and recovery log odds ratios in depression toy example

**Figure 13:**
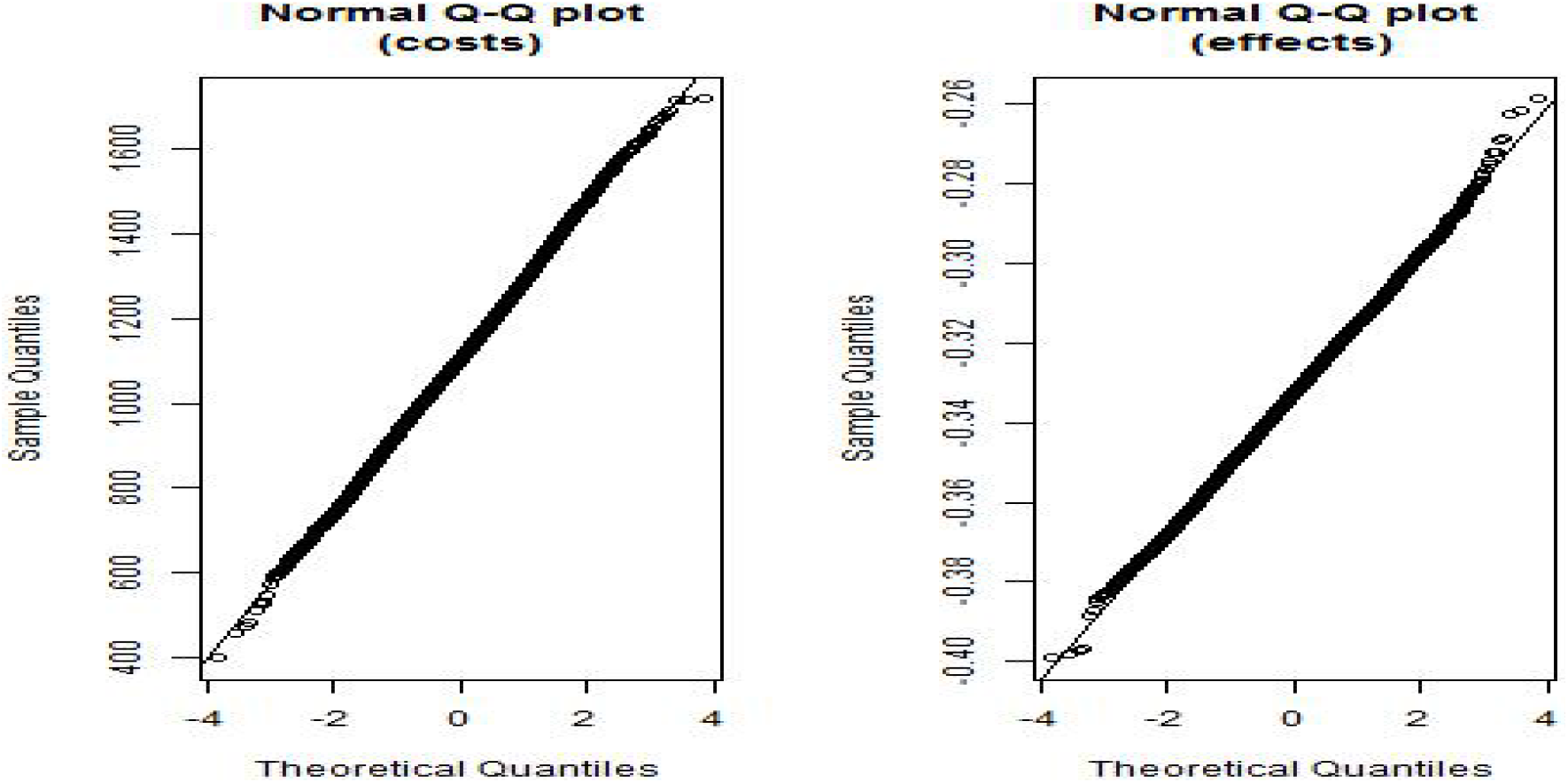
QQ-plots QQ-plots for INLA-GP estimate of EVPPI for simple trial parameter set in DOACs model

**Figure 14:**
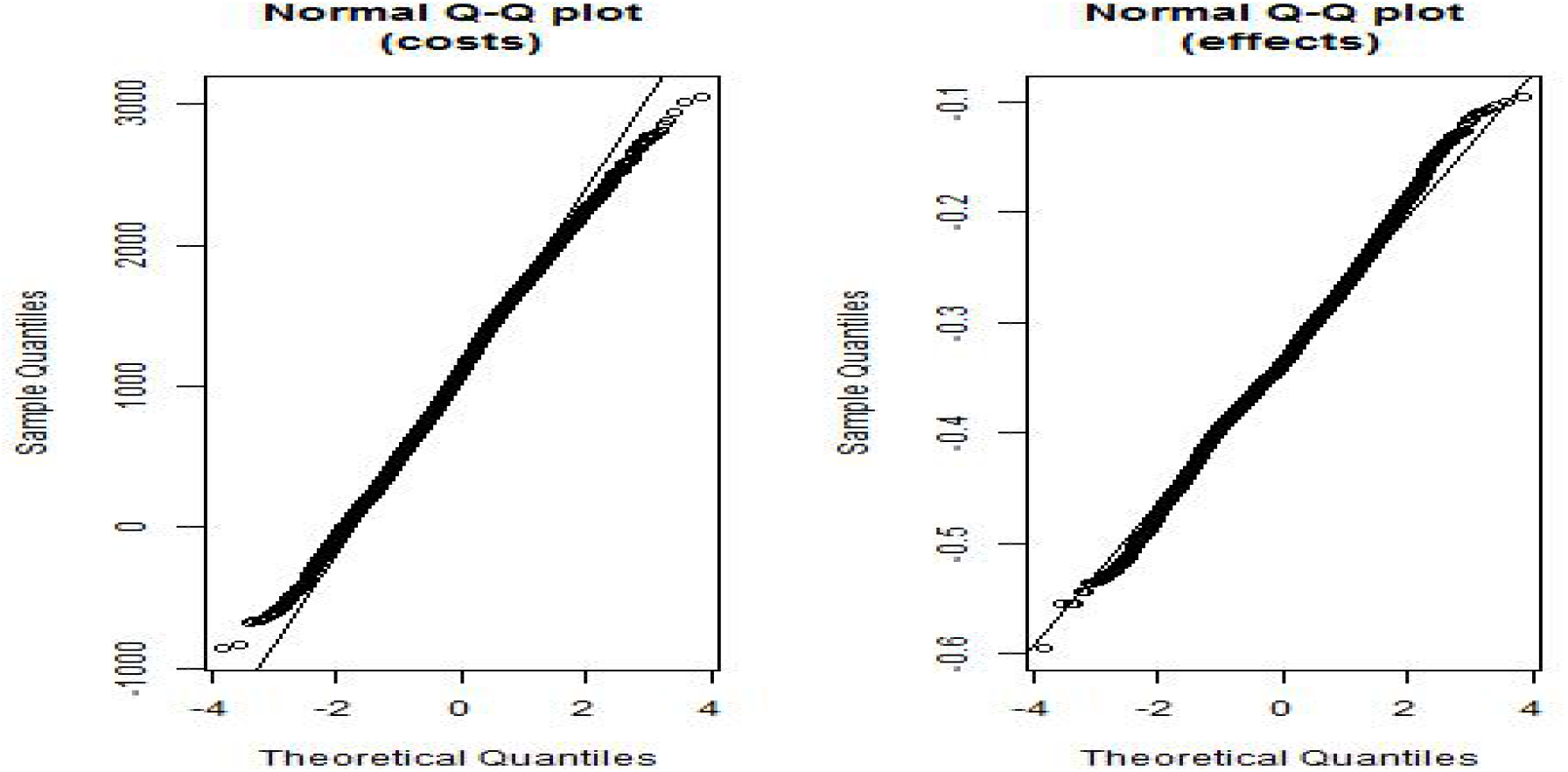
QQ-plots for INLA-GP estimate of EVPPI for log odds ratios in DOACs model

**Figure 15:**
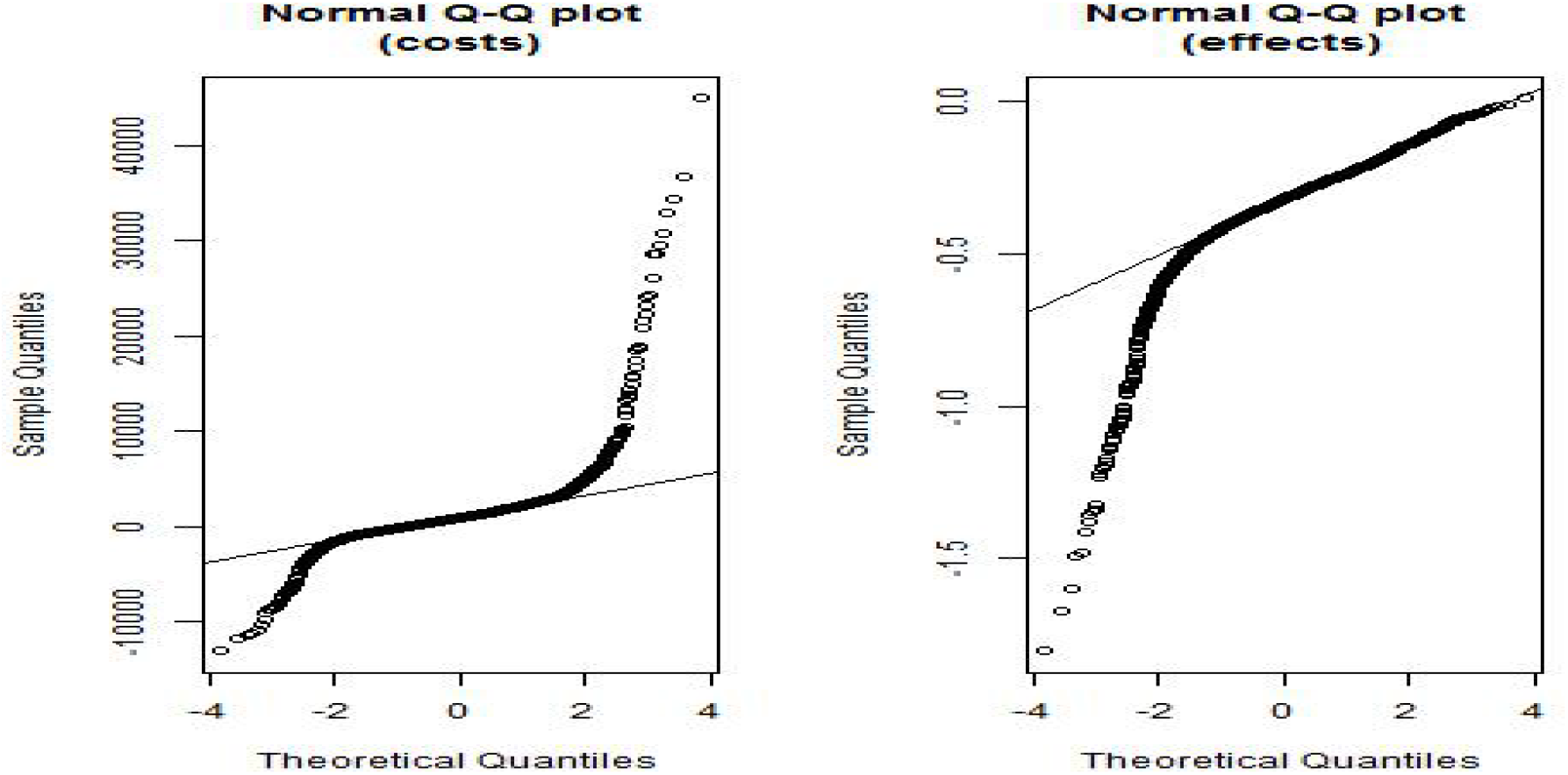
QQ-plots for INLA-GP estimate of EVPPI for all MCMC parameters in DOACs model

#### 7.3.3 EVPPI results of the 20 treatment depression model

For comparison, we set the MSE to be 0.25 (*ε* = 0.5). Table 10 shows the samples needed to achieve a 0.25 MSE. We observe that he computational cost of MC and MLMC in EVPPI for Cost and QALYs are much less than QMC. The reason is that MC and MLMC use DIFF estimator, whereas QMC directly calculates EVPPI. In this case, calculating DIFF is more efficient than EVPPI.

**Table 10:** Comparison of computational cost (measured as number of 10^6^ samples required to obtain an MSE of 0.25).

| EVPPI | Value | Credible interval | Computational Cost ($10^6$ samples) | | |
| --- | --- | --- | --- | --- | --- |
|  |  |  | MC | MLMC | QMC |
| $(P_{rec}^{(d)}, P_{rel}^{(d)})$ | 85.84 | [82.45, 89.23] | 13.18 | 27.41 | 0.32 |
| Cost and QALYs | 756.44 | [753.05, 759.83] | 0.46 | 0.23 | 3.2 |
| $lor_8$ and $lor_{10}$ | 83.68 | [80.29, 87.07] | 28.72 | 12.37 | 0.1 |

**Table 11:** EVPPI value 1024*128

|  | EVPPI | EVPPI sd | Bias | Credible interval |
| --- | --- | --- | --- | --- |
| Multivariate Normal | 184.51 | 16.01 | 3.85 | [152.23, 216.78] |
| Multivariate t distribution | 189.13 | 16.89 | 3.24 | [155.42, 222.83] |

### 7.3 Quantile-quantile diagnostic plots for BCEA INLA-GP EVPPI estimation

The qq-plots below were produced using the BCEA function **diag.evppi**.

### 7.4 Reference to GitHub repository with R code

We provide the in a supplemental file but it is also available, along with mlmc.R and mlmc.test.R, at https://github.com/Bogdasayen/mlmc_evppi_demo Full model and MLMC/QMC algorithm code are provided in the GitHub repository located at https://github.com/Bogdasayen/Example_MLMC_and_QMC_for_EVPPI We briefly point to relevant files below but refer to the repository for full details.

#### 7.4.1 MLMC code

The R implementation of the MLMC algorithm is available in the MLMC package. The main task for a user is to construct the EVPPI level estimator (7.1) or DIFF level estimator (**??**). The depression toy model described in Section 3.1 is provided in the repository `Bogdasayen/ Example_MLMC_and_QMC_for_EVPPI/ToyModel-MLMC` The “ReadMeFirst.pdf” file includes a full description of the code.

In addition the DOAC MLMC code is provided in the repository `Bogdasayen/Example_ MLMC_and_QMC_for_EVPPI/DOAC-MLMC` The model description and summary of the numerical results are in NOACS_math.pdf To run the MLMC code, please

1. Run `NOAC.AF.model.main.3.R` to load necessary files and data.
2. Run `MLMC_Core.R` to do a simple test locally.
3. Run `Parellel_Core.R` to run the code using parallel computing.

#### 7.4.2 QMC code (toy model)

To implement QMC estimate, we start from generating Sobal sequences for uniform distributed random variables *U*. Then, for a random variable *X* with cumulative distribution function (CDF) Φ, we get *X* by *X* = Φ^−1^(*U*). To calculate EVPPIs, we only need the inverse CDFs for normal distribution `qnorm.R` and *β* distrubution, which is implemented in `sfunc.R`^1^.

QMC for the depression toy model is provided in the repository `Bogdasayen/Example_MLMC_ and_QMC_for_EVPPI/ToyModel_QMC/` The “ReadMeFirst.pdf” file includes a full description of the code.

In addition, the DOAC QMC code is in the repository `Bogdasayen/Example_MLMC_and_QMC_for_EVPPI/DOAC_QMC/` Note: The only code needed for user to run is `NOAC_QMC_main.R` To run the QMC code, please

1. Open `NOAC_QMC_main.R`, change the baseline.directory
2. Run `NOAC_QMC_main.R`
3. See examples of running EVPPI at the end of `NOAC_QMC_main.R`

### 7.5 MVN vs MVT

The conventional version of the *p*-dimensional multivariate t (MVT) distribution *X* ∼ *t*_*p*_(*µ*, Σ, *ν*), with mean *µ*, scale matrix Σ (generally not the covariance of *X*), and degrees of freedom *ν* (which determine the thickness of the tail), has the probability density function

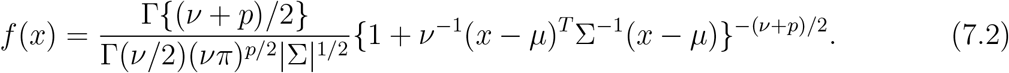

The conditional distribution of the multivariate t distribution also follows the multivariate t distribution. Assuming 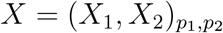, we can define

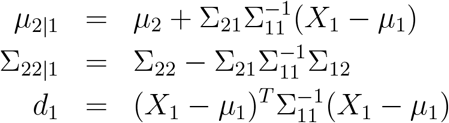

and then the conditional distribution of *X*_2_ given *X*_1_ is

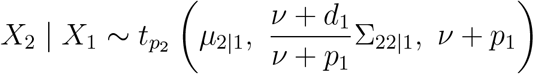

Compared with the multivariate normal distribution, the MVT can capture the feature of the thick tail and may give better approximation of the covariance structure of the MCMC samples. Here is the comparison results between the two distributions of the EVPPI for DOAC simple trial using standard MC with 1024 outer samples and 128 inner samples: from which we can see that the credible interval of MVT is consistent with the results when using multivariate normal distribution.

## Footnotes

1 This code is written by Thomas Lumley, for details see https://stat.ethz.ch/pipermail/r-help/2002-January/017624.html.

